# Initial antigen encounter determines robust T-cell immunity against SARS-CoV-2 BA.2.86 variant three years later

**DOI:** 10.1101/2024.08.09.24311705

**Authors:** Rocío Rubio, Alexei Yavlinsky, Marina Escalera Zamudio, Luis M Molinos-Albert, Carla Martín Pérez, Edwards Pradenas, Mar Canyelles, Cèlia Torres, Cedric Tan, Leo Swadling, Anna Ramírez-Morros, Benjamin Trinité, Josep Vidal-Alaball, Ruth Aguilar, Anna Ruiz-Comellas, Julià Blanco, Lucy van Dorp, François Balloux, Carlota Dobaño, Gemma Moncunill

**Affiliations:** ISGlobal, Barcelona, Spain; Facultat de Medicina i Ciències de la Salut, Universitat de Barcelona (UB), Barcelona, Spain; Institute of Health Informatics, University College London, London, UK; UCL Genetics Institute, University College London, London, UK; IrsiCaixa, Badalona, Spain; Division of Infection & Immunity, University College London, London, UK; Unitat de Suport a la Recerca de la Catalunya Central, Fundació Institut Universitari per a la recerca a l’Atenció Primària de Salut Jordi Gol i Gurina, Sant Fruitós de Bages, Spain; Health Promotion in Rural Areas Research Group, Gerència d’Atenció Primària i a la Comunitat Catalunya Central, Institut Català de la Salut, Sant Fruitós de Bages, Spain; Centre d’Atenció Primària (CAP) Sant Joan de Vilatorrada, Gerència d’Atenció Primària i a la Comunitat Catalunya Central, Institut Català de la Salut, Sant Fruitós de Bages, Spain; Institut Germans Trias I Pujol, IGTPO, Badalona, Spain; Universitat de Vic-Central de Catalunya, UVic-UCC, Vic, Spain; CIBER de Enfermedades Infecciosas (CIBERINFEC), Instituto de Salud Carlos III, Barcelona, Spain

**Author notes:** Shared corresponding authors.

**Keywords:** COVID-19, BA.2.86 cross-recognition, Pirola, T cells

## Abstract

**Background:** The emergence of the hypermutated severe acute respiratory syndrome coronavirus 2 (SARS-CoV-2) BA.2.86 variant raises significant concerns due to its potential to evade pre-existing immunity.

**Methods:** We measured cross-reactivity of neutralizing antibodies and T-cell responses to BA.2.86 in 52 previously exposed participants and investigated clinic-demographic and viral genetic determinants affecting T-cell responses.

**Findings:** We found that despite notable escape from neutralizing antibodies, T-cell responses remained generally preserved, albeit with a significant but small loss in T-cell cross-recognition (7·5 %, 14·2 % and 10·8 % average loss for IFN-γ, IL-2 and IFN-γ + IL-2, respectively, p < 0·05). This is consistent with the prediction of 6 out of 10 immunodominant T-cell epitopes (TCEs) altered by BA.2.86 lineage-defining mutations to have reduced peptide presentation. This effect is expected to be mitigated due to total TCEs across the genome. Remarkably, T-cell responses and cross-recognition were 3·5 (IFN-γ), 2 (IL-2) and 2·4 (IFN-γ + IL-2) times higher when first induced by natural infection rather than vaccination three years before, by increasing number of infections, and by ancestral/Delta than Omicron infections.

**Interpretation:** Our findings underscore the critical role and the factors influencing T-cell immunity against evolving SARS-CoV-2 variants, such as first antigen encounter (vaccination or infection), which is essential for developing effective control strategies against SARS-CoV-2 variants.

**Funding:** European Union (Horizon Europe), Fundació Privada Daniel Bravo Andreu, Catalan Government (PERIS, CERCA), Spanish Ministry of Science, Rosetrees Trust, Pears Foundation.

## Introduction

The evolution of severe acute respiratory syndrome coronavirus 2 (SARS-CoV-2) has led to the emergence and dominance of Omicron sublineages[1]. Currently (as of July 2024), the Omicron subvariant BA.2.86, also known as ‘Pirola’, and its immediate descendants, are the predominant variants circulating globally, accounting for over 99 % of SARS-CoV-2 cases[2].

This variant has raised significant concerns due to its 63 amino acid (aa) changes compared to the ancestral SARS-CoV-2 Wuhan spike (S) protein. These changes include 51 aa substitutions, 8 aa deletions, and 4 aa insertions[3]. The BA.2.86 variant exhibits a substantial genetic divergence from its predecessor, the BA.2 variant, with 38 aa changes in the S protein. This magnitude of change is comparable to the genetic leap observed between the Delta and Omicron variants[3,4]. BA.2.86 has evolved by acquiring convergent mutational sites that optimize the host receptor angiotensin converting enzyme 2 (ACE2) binding affinity, thereby enhancing infectivity and enabling immune evasion[3,5,6]. Although there is high vaccine coverage worldwide, these mutations may reduce vaccine effectiveness against infection, which has a significant socioeconomic impact on health[7].

Several recent studies have demonstrated that BA.2.86 exhibits extensive immune evasion from pre-existing humoral responses induced by vaccination, infection or any combination of both events[8–12]. However, few studies have examined the ability of S-specific T cells to cross-recognize BA.2.86 in-silico[4,13] or in-vivo[1,14].

Since it has been described that humoral responses elicited by current vaccines or SARS-CoV-2 infections are shorter-lived than T-cell responses[15], understanding the potential effects of viral mutations on cellular immune evasion is crucial for our knowledge of long-term immunity against SARS-CoV-2. Hence, we aimed to investigate the ability of BA.2.86 to escape pre-existing immunity, focusing particularly on T-cell responses and their determinants. To this end, we measured the cross-reactivity of neutralizing antibodies and T-cell responses in individuals previously exposed to viral infection and/or mRNA vaccination. Data were analysed in relation to clinical and sociodemographic characteristics. Additionally, we employed in-silico analysis to assess potential viral genetic determinants contributing to the differential T-cell responses and their impact on peptide binding affinity.

## Methods

### Study Design

Blood samples collected between May and June 2023 from 52 healthcare workers in the CovidCatCentral longitudinal cohort study created in 2020 were used to assess the evasion ability of the BA.2.86 variant from adaptive immune responses. The cohort is comprised of healthcare workers from several primary care counties in Barcelona, Spain[16–18]. The BA.2.86 variant was first documented in Spain on August 22^nd^, 2023. Plasma and cryopreserved peripheral blood mononuclear cells (PBMCs) from venous blood samples were used for neutralization and cellular assays, respectively. Sociodemographic and clinical information were recorded at each cross-sectional visit. SARS-CoV-2 asymptomatic or undiagnosed infections were identified by serology through fold change (FC) in antibody levels between timepoints. For participants vaccinated between timepoints, an individual was considered infected if the FC was greater than 4 for IgG or IgA against the nucleocapsid (N) antigen. For those not vaccinated between timepoints, an individual was considered infected if at least two antibody-antigen pairs among IgG and IgA against any of the S or N antigens had a FC ≥ 4 (Martin-Pérez C, in preparation). In the absence of sequencing data, we inferred probable variant infection based on the predominant viral variant circulating in Catalonia at the date of infection[19,20].

### Plasma Neutralizing Activity

Pseudovirus-based neutralization assay was performed using HIV reporter pseudoviruses expressing SARS-CoV-2 ancestral (WH1) and BA.2.86 S proteins and Luciferase gene, as previously reported[21]. The assay was performed in duplicate. Briefly, in 96-well cell culture plates (Thermo Fisher Scientific), 200 TCID50 of pseudovirus were preincubated with three-fold serial dilutions (1/60–1/14, 580) of heat-inactivated plasma samples at 37 °C for 1 h. Then, 1×10^4^ HEK293T/hACE2 cells treated with DEAE-Dextran (Sigma-Aldrich) were added. Results were read after 48 h using the EnSight Multimode Plate Reader and BriteLite Plus Luciferase reagent (PerkinElmer, USA). The values were normalized, and the ID50 (the reciprocal dilution inhibiting 50% of the infection) was calculated by plotting and fitting the log of plasma dilution versus response to a 4-parameters equation in Prism 10 (GraphPad Software, USA).

### Cellular Assay

The magnitude of the T-cell responses to the S protein from Wuhan and BA.2.86 was measured using the human IFN-γ/IL-2 FluoroSpot kit (Mabtech) as previously described[22].

The peptide pools used as stimulus included the full-length S protein from ancestral [PepTivator® SARS-CoV-2 Prot_S Complete (Miltenyi)] and from BA.2.86 [PepMix™ SARS-CoV-2 (Spike BA.2.86) (JPT)]. The peptides were 15 amino acids long with 11-amino acids overlaps and were dissolved in sterile water according to the manufacturer’s instructions.

PBMCs were isolated from venous blood samples by density-gradient centrifugation using Ficoll-Paque (Merck), cryopreserved in heat-inactivated fetal bovine serum (HI-FBS) (Thermo Fisher Scientific) with 10 % dimethyl sulfoxide (Merck), and stored in liquid nitrogen until use. After blocking the pre-coated FluoroSpot plates with culture medium-10% HI-FBS, 2·5×10^5^ thawed PBMCs were added to the stimulus (1 µg/mL/peptide concentration) or unstimulated control (only culture medium [TexMACS Medium (Miltenyi)-1% penicillin/streptomycin (Thermo Fisher Scientific)] wells, and 5×10^4^ PBMCs to the positive control (phytohemagglutinin (Merk), 5 µg/ml) wells. All conditions performed in duplicate, and were incubated at 37 °C and 5 % CO2 for 20 h.

Cells secreting IFN-γ and/or IL-2 were detected and counted as spot-forming units (SFU). Seven participants with ≥ 100 SFU in unstimulated wells for IFN-γ were excluded from the analysis.

SFU counts in the unstimulated wells were subtracted from those in the stimulated wells to account for background responses, and negative values were set to zero. The results were expressed as SFU / 10^6^ PBMCs. Responses were considered positive if the results were ≥ 3-fold the mean of their unstimulated wells for each cytokine and stimulus. Responders were defined as having a positive response to at least one cytokine-stimulus combination.

### Binding Antibody Assay

Luminex technology was used to measure binding IgM, IgG, and IgA levels (as median-fluorescence-intensity (MFI)) to the ancestral S, its subregions S2 and the RBD antigens from plasma samples as previously described[16].

### Genetic Determinants

Drawing from the complete 16·6 million SARS-CoV-2 genome sequence data available from GISAID[23], we filtered out genomes derived from non-human hosts, and those incomplete or with low-coverage. This resulted in a final dataset of 15 million sequences comprising a total of 27,503 mutations within S. Data was stratified according to virus lineage, aggregating data under the following criteria: 1) Earliest genome sequences (Wuhan-1 and those with collection dates before March 1^st^, 2020), 2) Pre-VOC lineages (genomes predating Alpha), 3-7) All VOCs: Alpha, Beta, Gamma, Delta and Omicron (each analysed separately), and 8) Variants of interest (VOI) Pirola (comprising the BA.2.86, JN.1, and descending sublineages). Heatmaps were generated in Python v3.8.10 using the matplotlib and seaborn packages. Each heatmap represents the normalised count score for the TCE within a given VOC/VOI stratum.

#### Normalised indel count scores

For each VOC/VOI, and for each CD4+ and CD8+ TCE, we counted the number sequences with indels affecting one or more sites at any position within a given TCE. We then divided count values by the total number of sequences belonging to each VOC/VOI, in order to obtain a normalised VOC/VOI indel score for each TCE.

#### Normalised substitution count scores

For each VOC/VOI, and for each CD4^+^ and CD8^+^ TCE, we considered only sequences in which the TCE was unaffected by indels. For each CD4^+^ TCE, we counted the number of sites affected by substitutions across the entire TCE. For each CD8^+^ TCE, we restricted substitution counts to only for anchor point binding to MHC class II (corresponding to positions 1-2,9-10 of each CD8+ TCE). For each VOC/VOI -TCE combination, we divided total count by the number of sequences considered.

#### In-silico predictions for mutation impacting epitope function

The impact of mutations within known T-cell epitopes was assessed using NetMHCpan[24] version 4.1 and NetMHCIIpan[25] version 4.1. The predicted binding of peptides corresponding to the original epitope sequence in Wuhan hu-1, the epitope sequence in BA.2.86 containing LDMs, or the epitope sequence in other lineages (e.g. JN.1) was calculated for the known MHC restriction. Where the MHC restriction was not known, HLA supertype representatives were used and included when weak or strong binding was predicted for the Wuhan hu-1 peptide sequence. A threshold of rank 0·5 % and 2·0 % were used to define strong and weak binders for MHC class I restricted epitopes and 1 % and 5·0 % for MHC class II restricted epitopes. Loss of an epitope due to reduced peptide-MHC binding was estimated when a peptide went from a strong binder to a weak, or a weak binder to a non-binder. Partial loss was defined as an increase in rank for the known restricting MHC and/or several predicted restricted MHCs of > 0·5 %.

### Ethics statement

The study protocol was approved by the IDIAP Jordi Gol Ethics committee (code 20/162-PCV) and written informed consent was obtained from all participants.

### Statistical Analyses

Sociodemographic and clinical data were compared between groups pf first antigen encounter using the CompareGroups R CRAN package[26]. For continuous normal variables, the mean and s.d. were calculated, and t-test were applied to assess differences. For continuous non-normal variables, the median and the first and third quartiles were calculated. For categorical variables, differences in proportions were calculated using chi-square test or Fisher’s exact test, when applicable.

Nonparametric tests were used to analyze neutralizing antibody and T-cell data. Nominal p-values of < 0·05 were considered statistically significant. We compared the neutralizing activity of plasma antibodies and S-specific T-cell responses between Wuhan and BA.2.86 using paired Wilcoxon Signed-Rank test. BA.2.86 recognition was assessed by calculating the FC in neutralizing activity and T-cell responses to BA.2.86 with respect to ancestral strain (BA.2.86 / ancestral). We compared the proportions (%) of secreting T cells induced by Wuhan vs. BA.2.86 strains using the chi-square test. Comparisons of magnitude of T-cell responses and BA.2.86 T cells recognition (estimated as BA.2.86/ancestral FC) between sociodemographic and clinical groups were performed by the Wilcoxon rank-sum test.

A multivariable linear regression model was fitted to assess the association between the magnitude of the T-cell responses to ancestral and BA.2.86 as the outcome variables and being infected prior to vaccination (first antigen encounter infection) as a predictor variable. This model was adjusted for three variables: the number of vaccine doses (continuous), total infections (continuous), and probable variant of infection (categorical). For the linear regression model, we checked the linearity of the data, normality of residuals, homogeneity of residual variance, independence of the residual error terms, and multicollinearity among the predictor variables. The models’ performance for ancestral and BA.2.86 had an Adjusted R^2^ of 0·37 and 0·36 for IFN-γ, 0·18 and 0·33 for IL-2 and 0·25 and 0·37 for IFN-γ + IL-2, respectively.

Correlations between antibody and cellular responses were assessed using Spearman’s rank correlation coefficient ρ (rho), and p-values were computed via the asymptotic t approximation. All data processing and statistical analyses were performed using R software version 4.2.3.

## Results

### Description of study population

We measured neutralizing antibodies and T-cell responses to Wuhan and BA.2.86 variant in blood samples from 52 healthcare workers participating in a prospective live COVID-19 cohort created in 2020, in blood samples collected between May and June 2023[16–18]. Clinic-demographic characteristics of participants are depicted in **Table 1**. The majority were female (85%) with an average age of 49 years (mean 49·17 s.d. 10.90). All participants had received three or four doses of an mRNA vaccine. The fourth dose was primarily (11/13) bivalent Original + Omicron BA.4/5. The median time since last vaccination was approximately 17 months. Among the participants, 49 had hybrid immunity and 3 had only vaccine-induced immunity. Forty-one individuals were likely infected with any of the Omicron subvariants due to the timing of infection, with a median time since last symptomatic infection of ∼ 16 months. Twenty-one participants had natural infection as their first antigen encounter (first-infected) and were then vaccinated, while 31 had vaccination as first exposure (first-vaccinated) (**Table 2**). Half of the participants reported at least one comorbidity, most participants were non-smoker, and none had long COVID.

**Table 1.**
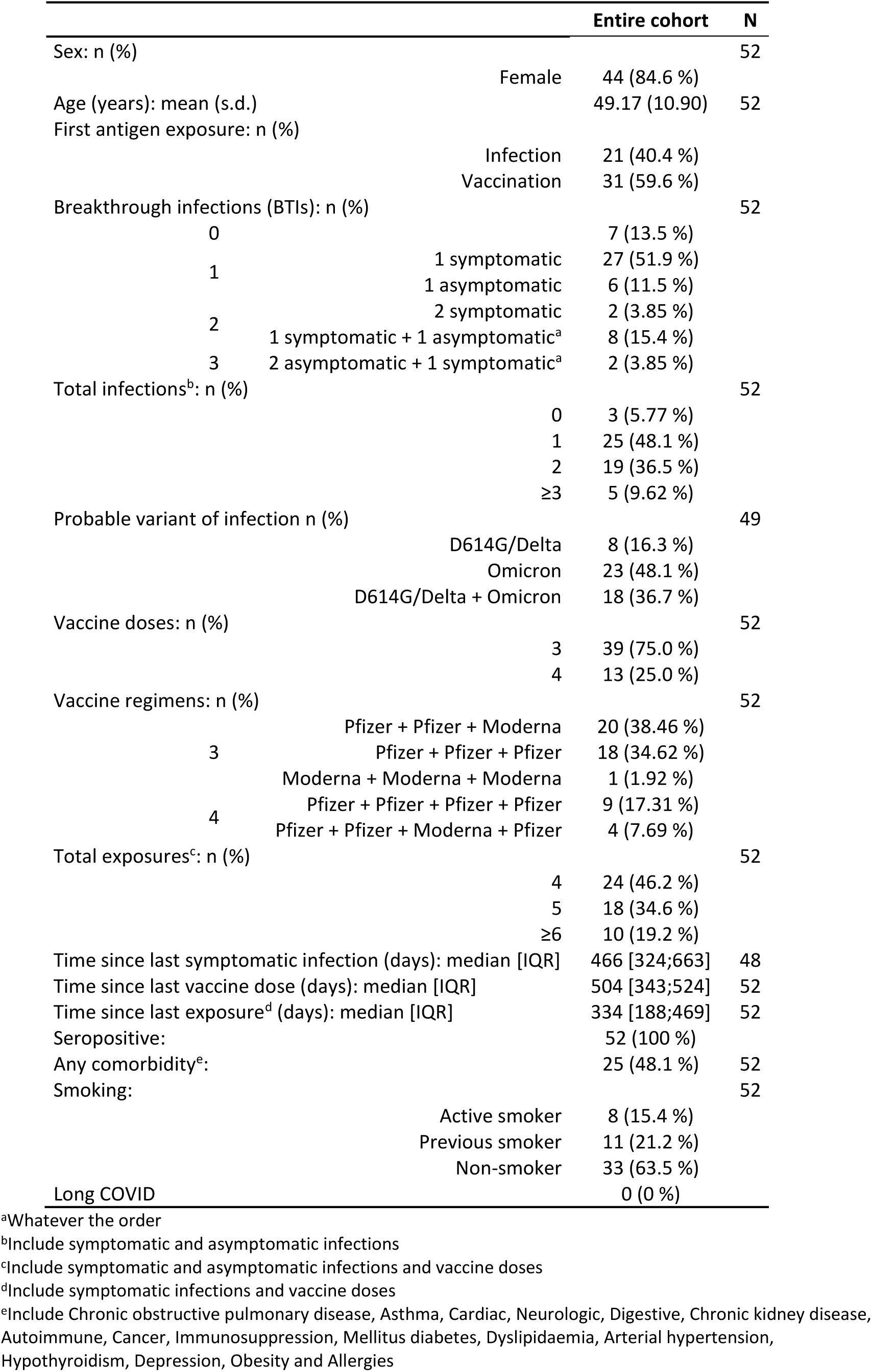
Sociodemographic and clinical characteristics of study participants.

**Table 2.**
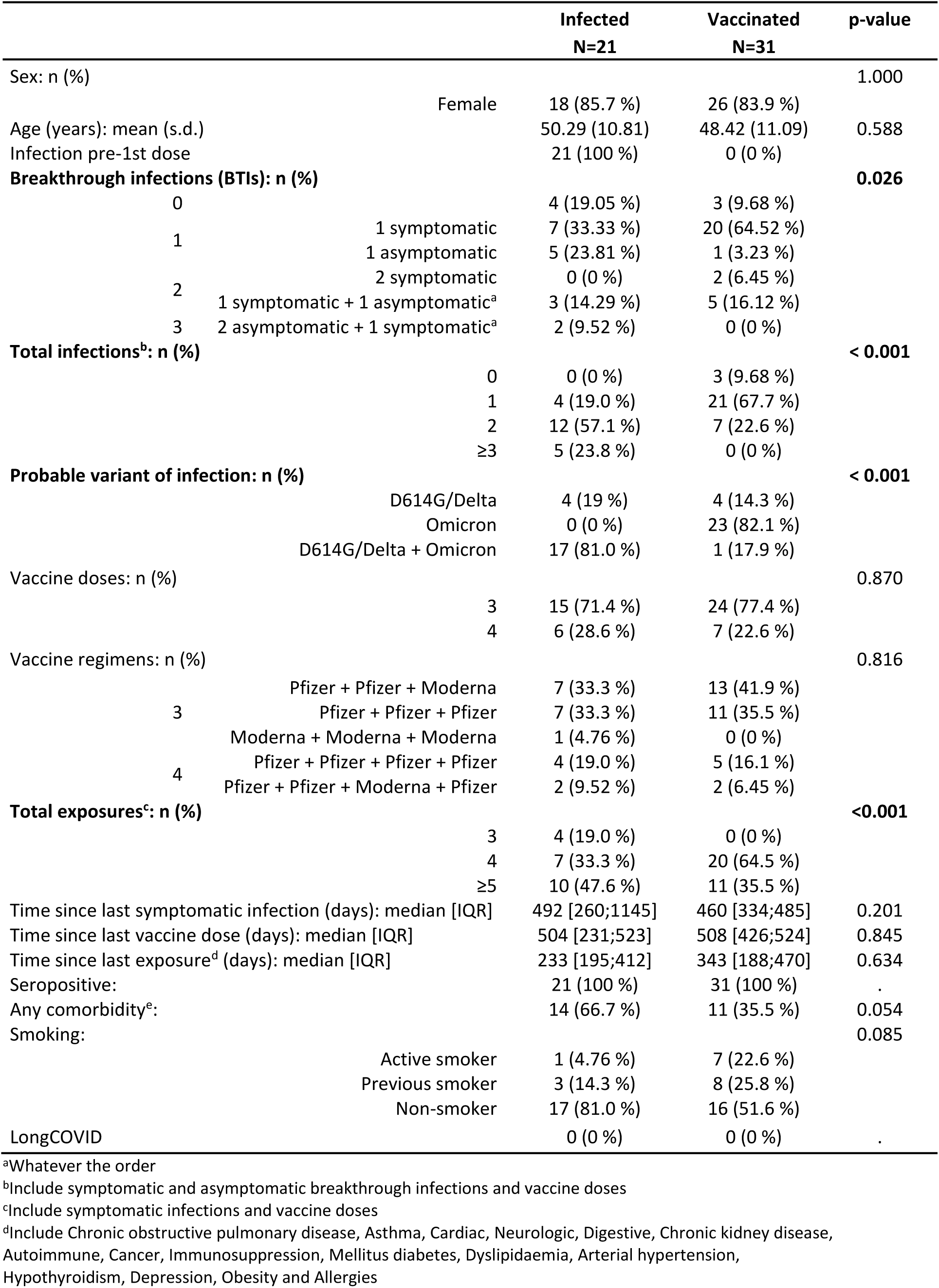
Sociodemographic and clinical characteristics by groups of first antigen encounter.

### Memory immune responses to Wuhan and BA.2.86

The neutralizing activity of plasma antibodies and S-specific T-cell responses to both Wuhan and BA.2.86 were measured using pseudovirus neutralization and IFN-γ/IL-2 FluoroSpot assays, respectively. After the last COVID-19 exposure (median 466, IQR: 188-469 days), with 85% of participants having been infected with the Omicron variant, the plasma neutralizing activity to BA.2.86 was significantly compromised (94·25 % average loss, p < 0.0001) compared to the Wuhan (**Fig. 1A**). In contrast, S-specific T-cell responses to BA.2.86 were significantly but only slightly reduced (7·5 %, 14·2 % and 10·8 % average loss for IFN-γ, IL-2 and IFN-γ + IL-2, respectively, p < 0·05) than those to ancestral strain (**Fig. 1B**). Additionally, T-cell responses to BA.2.86 strongly correlated with those to the Wuhan (rho = 0·91, p < 0·001, spearman test, **Fig. S1**). To quantify the BA.2.86 cross-recognition, we calculated the fold change (FC) of BA.2.86 responses relative to the ancestral strain (**Fig. 1C-D**). Despite BA.2.86 effectively evading a significant proportion of neutralizing antibodies, T-cell responses remained relatively intact.

**Fig. 1.**
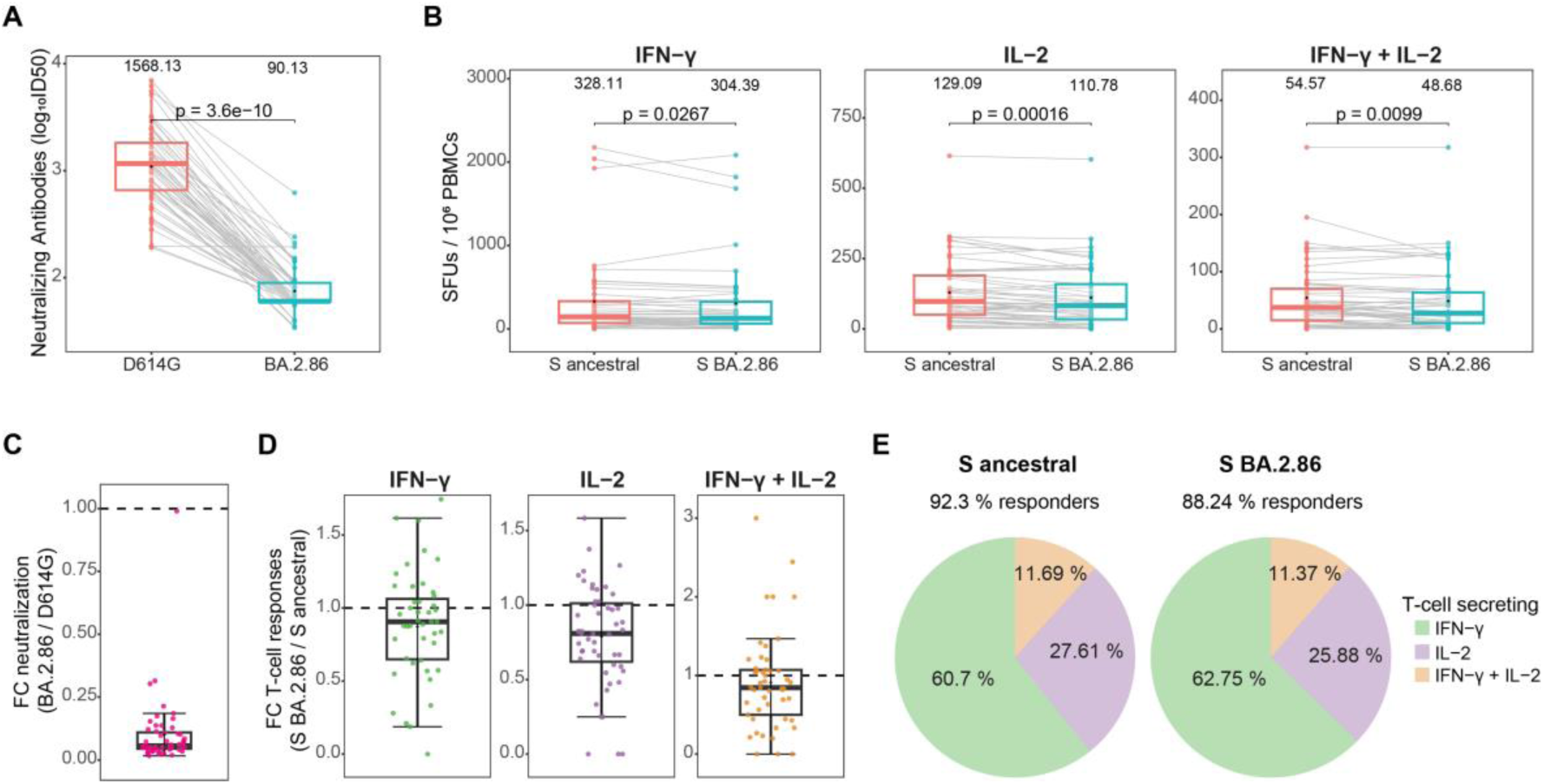
Neutralizing activity of plasma and T-cell responses to Wuhan and BA.2.86. Neutralizing activity of plasma antibodies expressed as log10ID50 (**A**) and S-specific T-cell responses as SFUs / 10^6^ peripheral blood mononuclear cells (PBMCs) secreting IFN-γ, IL-2 or IFN-γ + IL-2 (polyfunctional) to ancestral and BA.2.86 (**B**). Responses were compared by paired Wilcoxon Signed-Rank test. Mean values are on the top. Fold change (FC) in neutralizing activity (**C**) and T-cell responses (**D**) to BA.2.86 with respect to ancestral strain (BA.2.86 / ancestral). Boxplots represent median (bold line), the mean (black diamond), 1^st^ and 3^rd^ quartiles (box), and largest and smallest values within 1.5 times the interquartile range (whiskers). (**E**) Percentage of responders to each viral strains and pie charts showing the average proportion (%) of secreting T cells by cytokine: IFN-γ (green), IL-2 (purple), or both (orange) by viral strain. Proportions were compared by Chi-square test, and there were not statistically significant differences. Interferon-gamma (IFN-γ), interleukin-2 (IL-2), spike (S), spot-forming units (SFU), D614G (ancestral). Sample size was 52 except for T-cells secreting IFN-γ (N = 45) and BA.2.86-specific T cells secreting IL-2 or IFN-γ + IL-2 (N = 51).

Nearly all participants (92·31%) had detectable T-cell responses to Wuhan, with a slight decrease in responders (88·24%) observed for BA.2.86 (**Fig. 1E**). T-cell responses to Wuhan and BA.2.86 predominantly secreted IFN-γ (61 and 63 %, respectively), followed by IL-2 (28 and 26 %), with a minority secreting IFN-γ + IL-2 (12 and 11 %), indicative of polyfunctionality (**Fig.1E**).

Correlations between plasma neutralizing activity and T-cell responses for Wuhan or BA.2.86 were notably weak (rho ranging -0·002 and 0·22, p > 0·05, spearman test, **Fig. S2**).

### Clinic-demographic factors influencing T-cell responses to BA.2.86

We aimed to elucidate the clinic-demographic factors associated to S BA.2.86 recognition by T cells. No associations in the T-cell responses to BA.2.86 or cross-recognition (FC BA.2.86/ancestral) were found in relation to sex (p > 0·05, **Fig. S3**), presence of any comorbidity (p < 0·05, **Fig. S4**), smoking status (p > 0·05, **Fig. S5**), immunity groups (hybrid immunity vs. only vaccinated) (p > 0·05, **Fig. S6**), number of vaccine doses (p > 0·05, **Fig. S7**), nor total number of exposures (including both vaccine doses and infections) (p > 0·05, **Fig. S8**).

#### Decreased T-cell responses to BA.2.86 by Omicron infection

While no differences in the magnitude of T-cell responses to BA.2.86 were found based on the number of infections (p > 0·05, **Fig. S9**), individuals who had experienced only one infection exhibited decreased recognition of BA.2.86 by IL-2 and polyfunctional secreting T cells compared to participants infected twice (**Fig. 2A**). Subsequently, we investigated which variants were responsible for influencing BA.2.86 T-cell recognition. In the absence of sequencing data, we inferred probable variant infection based on the predominant viral variant circulating in Catalonia at the date of infection[19,20]. Participants infected by earlier variants, ancestral or Delta, exhibited a greater magnitude of T-cell responses to Wuhan and BA.2.86 compared to participants infected with Omicron variants, although differences only reached statistical significance for the Wuhan (**Fig. 2B**). Furthermore, individuals infected with only Omicron variants showed a decreased magnitude of T-cell responses (by IFN-γ and polyfunctional T cells) to BA.2.86 (**Fig. 2B**), and decreased BA.2.86 recognition by IL-2 secreting T cells (**Fig. 2C**) than individuals infected with both earlier strains (ancestral or Delta) and Omicron.

**Fig. 2.**
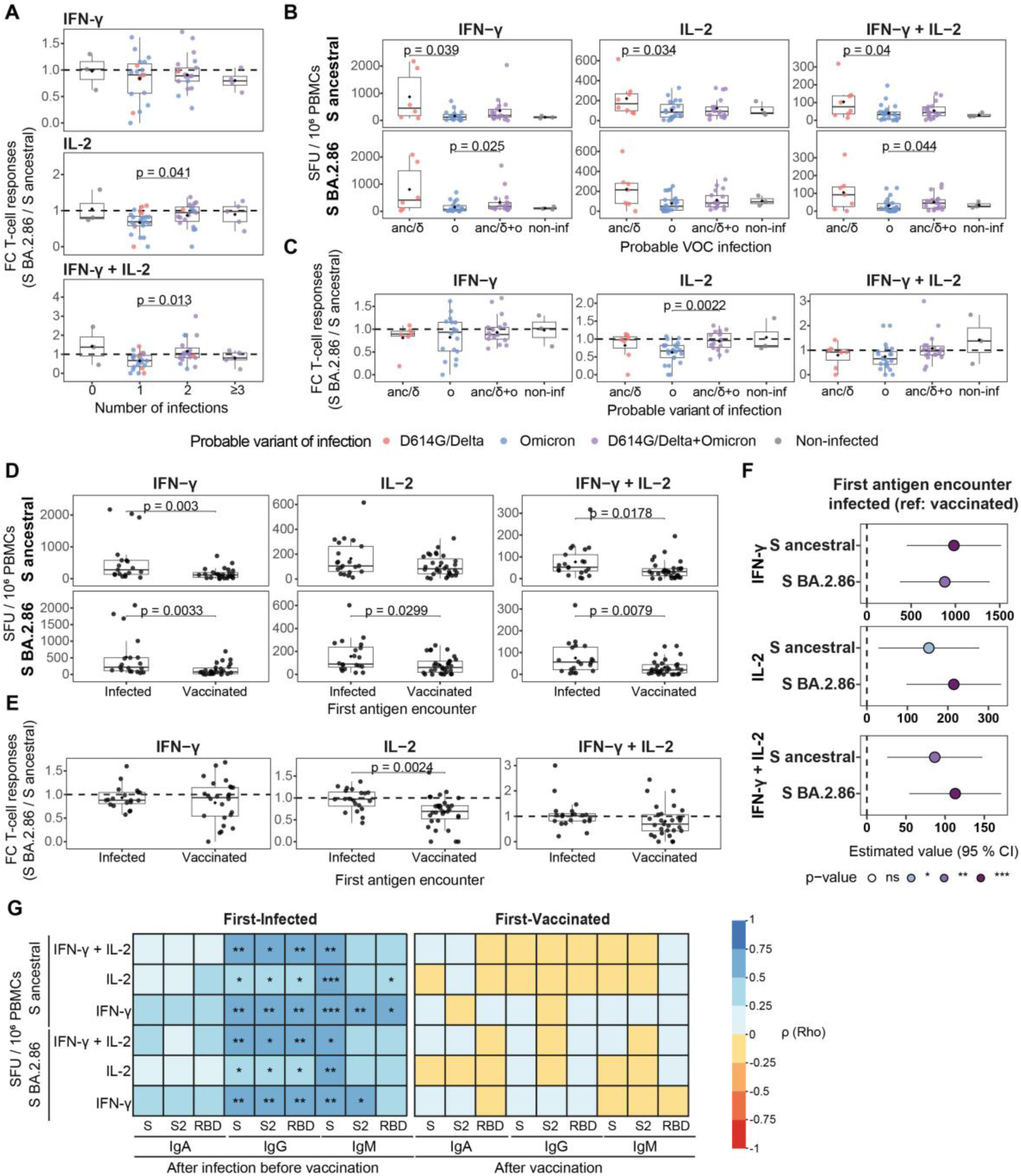
Factors influencing T-cell responses to ancestral and BA.2.86 spike. (**A**) BA.2.86 recognition as FC in T-cell responses to BA.2.86 with respect to ancestral strain (BA.2.86 / ancestral) by number of SARS-CoV-2 infections. (**B**) Magnitude of T-cell responses as SFU / 10^6^ peripheral blood mononuclear cells (PBMCs) of T-cells secreting IFN-γ, IL-2 or IFN-γ + IL-2 (polyfunctional) and (**C**) BA.2.86 T-cell recognition by probable variant of infection. (**D**) Magnitude of T-cell responses and (**E**) BA.2.86 recognition by first antigen encounter. T-cell responses were compared by Wilcoxon rank-sum test. Boxplots represent median (bold line), the mean (black diamond), 1^st^ and 3^rd^ quartiles (box), and largest and smallest values within 1.5 times the interquartile range (whiskers). (**F**) Forest plot showing the association of being infected before vaccination with magnitude of T-cell responses. The represented values and CI show the SFU / 10^6^ PBMCs increase in individuals infected before vaccination compared to individuals not infected prior vaccination Multivariable linear regression models were fitted to calculate the estimates (dots) and 95 % confidence intervals (lines) and adjusted for the number of vaccine doses, total infections and the probable variant of infection. (**G**) Heatmaps illustrating the Spearman’s correlation coefficient ρ (Rho) between the magnitude of T-cell responses three years post-first infection or 28 months post-vaccination with the antibody responses (median fluorescence intensity (MFI) of IgA, IgG, and IgM) to S, S2 and RBD from ancestral strain 5 months post-first infection (first-infected) and 3 months post-first vaccination (first-vaccinated). p-values: * ≤ 0.05, ** ≤ 0.01 and *** ≤ 0.001. Anc/δ (ancestral/Delta), ο (Omicron), anc/δ + ο (ancestral/Delta + Omicron), non-inf (non-infected). Interferon-gamma (IFN-γ), interleukin-2 (IL-2), spike (S), receptor binding domain (RBD), spot-forming units (SFU). Sample sizes by number of infections: 0 N = 3, 1 N = 24, 2 N = 19, ≥ 3 N = 5; by probable variant: ancestral/Delta N = 8, Omicron N = 23, D614G/Delta+Omicron N = 18; by first antigen encounter: Infected N = 21, Vaccinated N = 31.

#### Increased T-cell responses to BA.2.86 by infection before vaccination 3 years earlier

We found that participants who had been infected before vaccination (first-infected) showed an increased magnitude of T-cell responses three years later to both ancestral (3·5 and 1·9 times higher for IFN-γ and IFN-γ + IL-2, respectively) and BA.2.86 (3·5, 2 and 2·4 times higher for IFN-γ, IL-2 and IFN-γ + IL-2, respectively) compared to participants without infection before vaccination (first-vaccinated) (**Fig. 2D**). Additionally, first-infected participants exhibited 1·5 higher BA.2.86 cross-recognition by IL-2 secreting T cells (**Fig. 2E**). After adjusting in multivariable linear regression models for the potential confounders (number of vaccine doses, total infections, and probable variant) (**Table 2**), infection before vaccination was still significantly associated with increased magnitude of T-cell responses to Wuhan and BA.2.86 strains 3 years after exposure compared to individuals who were first-vaccinated (**Fig. 2F**).

Furthermore, in the first-infected group, the magnitude of T-cell responses to both Wuhan and BA.2.86 three years after the first SARS-CoV-2 infection was positively correlated with the binding antibody levels measured 5 months (mean 151·7, IQR 60·5 (days)) after first infection (**Fig. 2G**), especially to IgG (rho for ancestral: IFN-γ 0·66, IL-2 0·51, IFN-γ + IL-2 0·59, p < 0·05; rho for BA.2.86: IFN-γ 0·66, IL-2 0·52, IFN-γ + IL-2 0·59, p < 0·05) and IgM anti-S antigens (rho for Wuhan: IFN-γ 0·74, IL-2 0·72, IFN-γ + IL-2 0·63, p < 0·01; rho for BA.2.86: IFN-γ 0·69, IL-2 0·67, IFN-γ + IL-2 0·57, p < 0·05, spearman test). Conversely, in the first-vaccinated group, the magnitude of T-cell responses 28 months after primary vaccination (January 2021) was not correlated with any of IgG, IgA and IgM levels measured 3 months (mean 93·42, IQR 11 (days)) after primary vaccination (rho < 0·12, p > 0·05, spearman test **Fig. 2G**). Moreover, there was no correlation between the magnitude of T-cell responses to any of the two lineages and the antibody levels measured at the same time point in the first-vaccinated or first-infected group, except for IgG and IgM anti-S2 in the first-infected group (rho ≈ 0·54, p < 0·05, spearman test, **Fig. S10**).

### Genomic correlates of T-cell responses

We then investigated potential genetic determinants as contributors to the differential T-cell responses observed to BA.2.86 compared to other virus lineages. Through an in-silico analysis of a comprehensive dataset comprising approximately 16 million SARS-CoV-2 genomes available from GISAID[23,27], we computed mutation frequencies for indels (insertions or deletions) and point substitutions within the S protein, normalized by the total number of sequences per virus lineage. Assuming that aa changes within T-cell epitopes (TCEs) may lead to a reduction or loss of epitope binding/recognition, we mapped the mutations onto the S protein sequence to identify those falling within immunodominant CD8^+^ and CD4^+^ TCEs reported [28]. This list of S mutations represents the most comprehensive TCE data associated with SARS-CoV-2 available to date (**Tables S1 and S2**). We considered substitutions and indels separately, as the latter are expected to have a stronger impact on epitope function (e.g., full disruption of recognition sites).

#### Lineage-specific patterns of mutations within T-cell epitopes

Immunodominant CD8^+^ (n=41) and CD4^+^ (n=55) TCEs locate within the S1 and S2 protein domains (**Tables S1 and S2**). An apparent lineage-specific pattern for indels and substitutions was observed for both CD8^+^ and CD4^+^ TCEs, in which Omicron (defined as the B.1.1.529 and descending sublineages prior to the emergence of the BA.2.86) and BA.2.86 share similar profiles (**Fig. 3**).

**Fig. 3.**
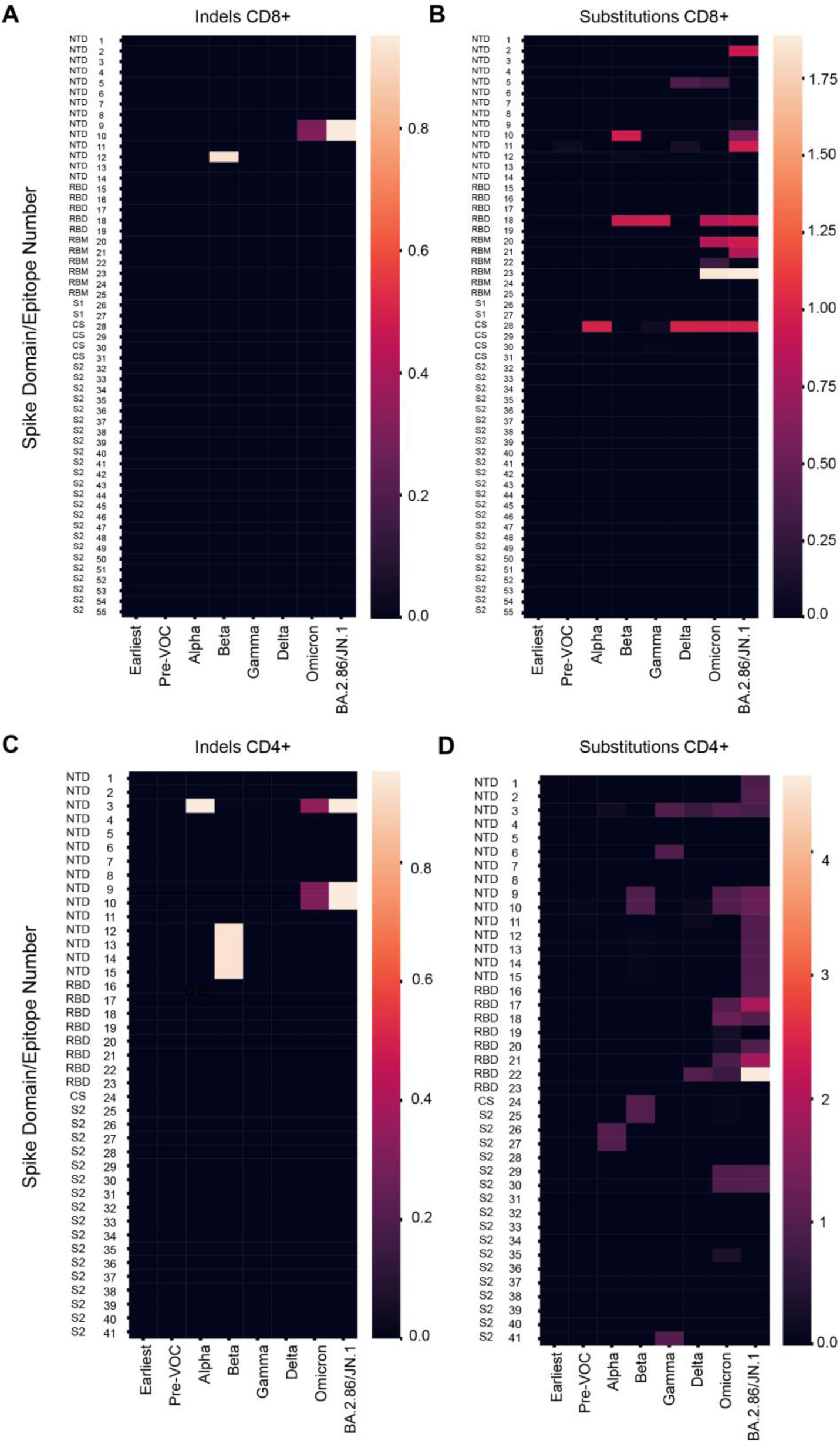
Mutation frequencies within the S protein on T-cell epitopes (TCEs) by SARS-CoV-2 lineages. Heatmaps showing mutation frequencies, indels and substitutions, in immunodominant CD8^+^ (**A-B**) and CD4^+^ (**C-D**) TCEs across different SARS-CoV-2 variants. Earliest genome sequences include Wuhan-1 and those with collection dates before March 1^st^, 2020, Pre-VOC lineages include genomes predating Alpha variant.

For indels, CD8^+^ TCEs epitopes 9 and 10 falling within the N-terminal domain (NTD) of S1 were affected (**Fig. 3A**). For CD4^+^ TCEs, epitopes 3, 9, and 10, also falling within the NTD of S1, were affected (**Fig. 3C**). Although affected TCEs were the same between Omicron and the BA.2.86 variant, average indel counts in the BA.2.86 were approximately 3-fold higher than in Omicron (3·17-fold for epitope 9 and 3·12-fold for epitope 10 in CD8^+^ and 2·75-fold for epitope 3 and 3·12 for epitopes 9 and 10 in CD4^+^ TCEs).

For substitutions, a higher proportion of CD8^+^ and CD4^+^ TCEs were found to be affected (**Fig. 3B and D**), consistent with these being better tolerated at the protein function level. Again, Omicron and BA.2.86 shared a similar profile, with an increased number of substitutions observed in BA.2.86 consistent with genetic divergence. For CD8^+^ TCEs, Omicron showed 6 affected epitopes, while the BA.2.86 showed 8. From these, 4 were overlapping between lineages, and 4 showed increased substitution counts in the BA.2.86, with epitopes 2, 10, 11, 20 and 21 being affected (**Fig. 3B**). For CD4^+^ TCEs, Omicron showed 10 affected epitopes, while the BA.2.86 showed 18. From these, 9 were overlapping between lineages, and 16 showed increased substitution counts in the BA.2.86, with epitopes 1, 2, 9-17, 20, 21 and 22 being affected (**Fig. 3D**). Relative to pre-variants of concern (VOC) virus lineages, all CD8^+^ and CD4^+^ TCEs affected by either indels or substitutions in BA.2.86 are derived (novel), and expected to yield a loss or reduction in epitope binding/recognition by T cells.

#### BA.2.86-specific substitutions are predicted to affect peptide-MHC binding

When further tracking specific aa changes falling within CD8^+^ and CD4^+^ TCEs, we found a subset of 14 BA.2.86 (and JN.1)-specific lineage defining mutations (LDMs) potentially impacting epitope function: S50L, V127F, L216F, H245N, I332V, D339H, K356T, V445H, G446S, N450D, L452W, N460K, L455S and H681R. For LDMs affecting only CD8^+^ TCEs, S50L falls in epitope 2, L216F in epitopes 10 and 11, H245N in epitope 12, G446S in epitope 20, N450D and L452W in epitope 21, L455S in epitope 21 (with L455S being unique to the JN.1) and H681R in epitope 28. For LDMs affecting only CD4^+^ TCEs, S50L falls in epitope 1, V127F in epitope 2, L216F in epitope 11, H245N in epitopes 12, 14, and 15, I332V in epitope 16 and 17, D339H in epitopes 17 and 18, K356T in epitope 20, V445H in epitope 21, N460K in epitope 22 and L455S in epitope 21 (with L455S being unique to the JN.1). CD4^+^ TCEs 9 and 10 and CD8^+^ TCE 10 are affected by the non-LDM deletion at position 211. Following in-silico analyses predicting the impact of mutations on peptide binding affinity to major histocompatibility complexes (MHCs), we identified CD8^+^ TCEs 2, 10, 12, and 28 as potentially affected, with changes to epitopes 10, 12 and 28 being predicted to result in a significant loss of the likelihood for peptide presentation driven by specific LDM (**Table 3**). Due to limitations in estimating peptide binding to MHC class II or unknown restriction, for CD4^+^ TCEs, only epitopes 16, 20, and 22 were predicted to be human leukocyte antigen (HLA) binders of these previously described epitopes; however, BA.2.86 LDMs were predicted to strongly affect both epitopes 20 and 22 (**Table 4**). In summary, LDMs acquired by BA.2.86 are predicted to affect its recognition at several immunodominant CD4^+^ and CD8^+^ TCEs.

**Table 3.**
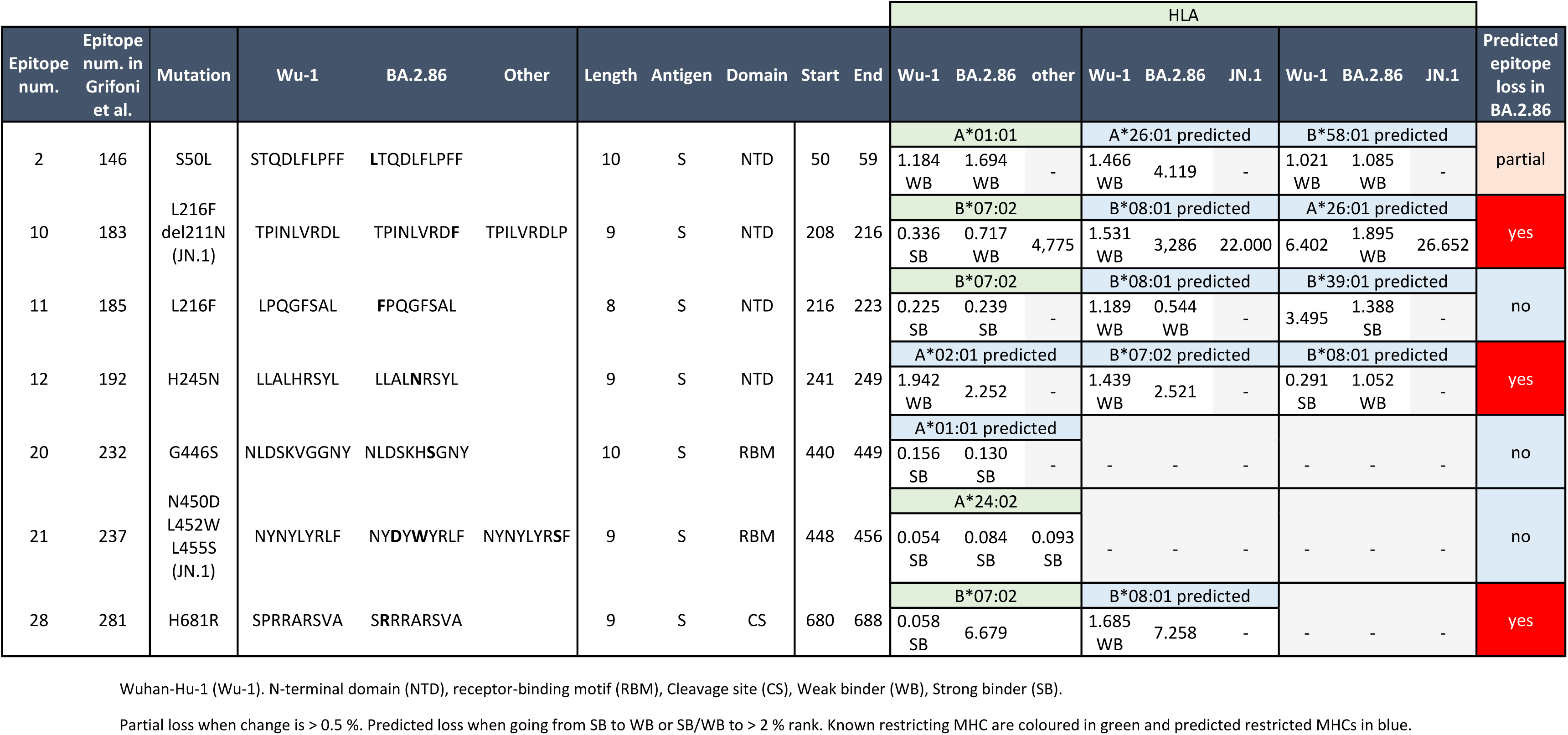
In-silico prediction of CD8^+^ T-cell epitope loss in BA.2.86 SARS-CoV-2 spike protein.

**Table 4.**
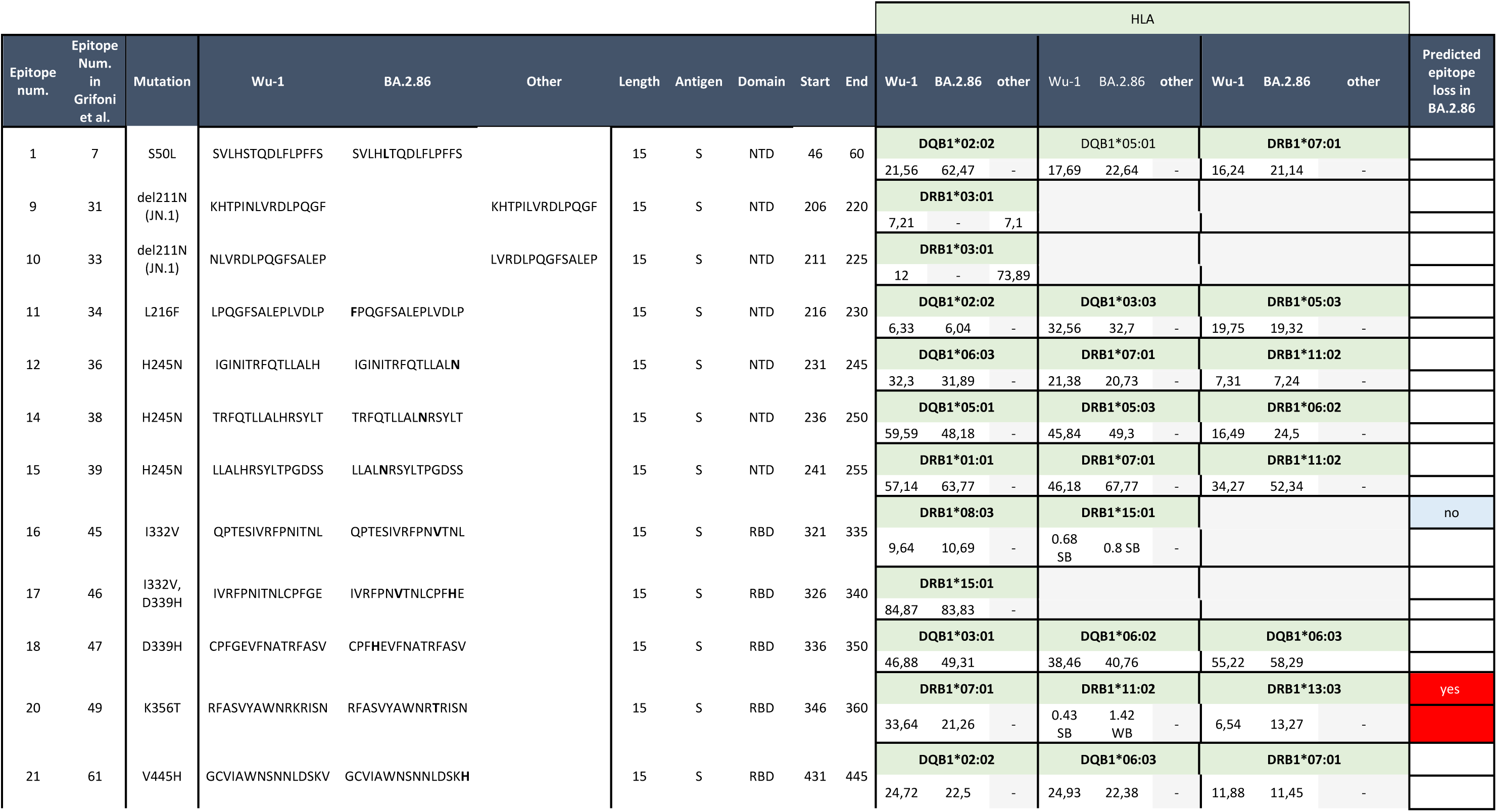
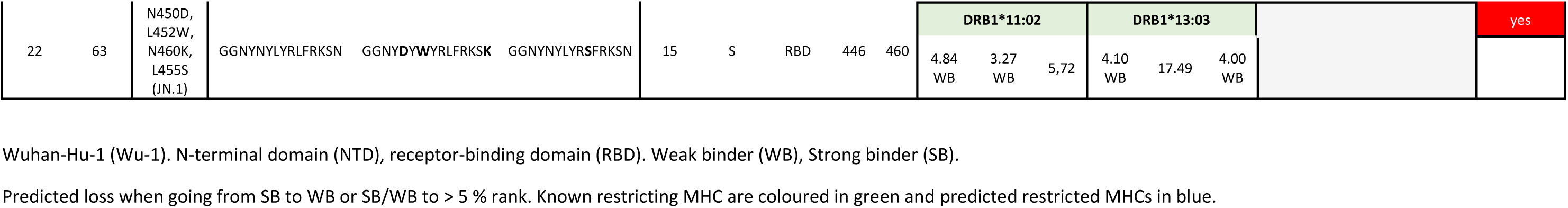
In-silico prediction of CD4^+^ T-cell epitope loss in BA.2.86 SARS-CoV-2 spike protein.

## Discussion

Despite the notable escape of BA.2.86 from pre-existing humoral immunity, T-cell responses remained, in general, preserved in individuals previously exposed through vaccination and/or infection. Furthermore, although a considerable proportion of LDMs are expected to affect BA.2.86-specific epitope function, with at least five TCEs predicted to be (total) lost, the overall impact is expected to be mitigated due to the majority of TCEs being still unaffected by mutations, translating into the minimal effect observed in functional assays. Moreover, TCEs located in S antigen are only a subset of all TCEs distributed across the viral genome. The large number of TCEs and their higher conservation outside S precludes effective T-cell immune evasion in previously infected individuals. Nevertheless, and of interest, our analysis revealed that T-cell responses and cross-recognition of BA.2.86 were heterogeneous in our study population, and were influenced by various factors, including the number of infections, the specific variant encountered, and the nature of the first antigen exposure (whether through vaccination or infection) despite 3 years had passed since then and with additional vaccine and infection exposures.

Our results confirm prior research[8–11], in which BA.2.86 exhibited substantial immune evasion from pre-existing neutralizing antibodies. However, T-cell responses were relatively well-preserved, consistent with the findings from limited studies on the cross-recognition of BA.2.86[1,14] and other variants[1,29–31]. Our data shows that T cells responding to both Wuhan and BA.2.86 predominantly secreted IFN-γ, followed by IL-2, with a minority of polyfunctional cells, as previously described for SARS-CoV-2[32]. Notably, these T-cell responses did not correlate with binding antibody levels nor neutralizing activity, indicating a discrepancy between antibodies and T cell-mediated immunity in terms of variant cross-recognition[33]. Viral selected mutations act as a double edge-sword by enhancing infectivity and evading humoral immune responses[3]. The consistent preservation of T-cell responses across variants suggests that most targeted epitopes are located in stable regions of the S protein, or that the mutations do not impair epitope recognition[1]. This preservation of S-specific T-cell responses underscores their potential importance in a context of declining neutralizing antibody responses against successively evolving variants[1]. Moreover, the lesser number of mutations in non-S proteins compared to S proteins could lead to broader and more robust T-cell variant recognition in previously infected individuals.

One might expect that two infections, particularly if at least one involves an Omicron variant, would lead to greater BA.2.86 cross-recognition by T cells compared to a single infection in the context of mild-to-moderate infections[1,34,35]. While we confirmed this, we found that being infected solely by Omicron variants, regardless the number of infections, decreased the magnitude of the T-cell responses and the recognition of BA.2.86 compared to being infected by ancestral/Delta or ancestral/Delta + Omicron. This observation might be explained by the combination of accelerated antigen clearance due to pre-existing immunity[36] and/or attenuated severity of Omicron variants; although reinfection rates during the Omicron epidemic were higher than in previous epidemic periods, the symptoms and infectivity have been observed to be milder than those of prior infections[37–39].

Remarkably, we observed that first-infected participants, displayed stronger T-cell responses three years later to both Wuhan and BA.2.86, as well as higher BA.2.86 recognition, compared to participants whose first antigen encounter was through vaccination (first-vaccinated). Since in the first-infected group all infections were mild-to-moderate, we used anti-S binding antibody responses as a proxy for the magnitude of infection to assess its association with T-cell responses after three years. T-cell responses were positively correlated with antibody levels after infection. In contrast, in the first-vaccinated group, T-cell responses did not correlate with antibody levels after primary vaccination. In addition, when comparing to the ancestral Wuhan-Hu-1 (vaccine) strain or early pre-VOC viral lineages, we found no significant enrichment of mutations suggesting that this was not the cause of the differential T-cell responses between the vaccine strain and early infection variants. Thus, our results suggest that instead of a mutation-driven immune priming process, exposure to the whole virus (offering a wider repertoire of antigens) and a stronger immune response after the first encounter might shape a more robust and sustained T-cell immune response. Supporting our hypothesis, previous studies have reported that initial COVID-19 severity imprints the long-term maintenance of SARS-CoV-2 adaptive immunity, with severe cases exhibiting more sustained virus-specific antibodies and memory T-cell responses compared to mild/moderate counterparts[40]. In parallel with our results, a previous study observed differences in transcriptional profiles and epigenetic landscape of S-specific CD4^+^ T cells between infected and vaccine-primed individuals two years after the encounter, with the infection-primed group showing enrichment for transcripts related to cytotoxicity and IFN-stimulated genes[41].

Additionally, other studies have reported higher T-cell responses over time in first-infected individuals[42,43], as well as higher frequencies of atypical memory B cell subsets and TH1 polarization of S-specific follicular helper T cells[44]. These findings warrant further investigation.

Finally, although a significant proportion of LDMs affect TCEs, the emergence of LDMs is not expected to be driven by selective forces exerted by T-cell immunity. LDM emergence and fixation may be driven by multiple evolutionary processes, including genetic drift (chance), or overlapping functional properties, such as ACE2 binding and cleavage for those TCEs falling within the receptor-binding motif (RBM) or cleavage site (CS) of S. Congruent with this, most mutations (substitutions/indels) affecting TCEs occur within the NTD and receptor-binding domain (RBD) of S1. In contrast, few mutations affecting TCEs fall within S2, largely reflecting a high degree of protein conservation across coronaviruses, which suggests less tolerance to changes given the high functional constraint.

Our study is limited by its relatively small sample size and most of the participants were female, which restricts the generalizability of our findings to broader populations such as older or immunocompromised individuals. Also, we have measured the magnitude of T-cell responses through FluoroSpot which cannot differentiate between CD4^+^ and CD8^+^ T-cell responses.

Overall, we report cross-reactive T-cell responses to the BA.2.86 variant in previously exposed healthcare workers, identifying the type of first-exposure as one relevant determinant of the extent of this cross-recognition. Our findings highlight the importance of T-cell immunity against evolving SARS-CoV-2 variants as key to counteract the notable escape from neutralizing antibodies. This suggests that T-cell immunity could be a crucial target for next-generation COVID-19 vaccines. Additionally, the history of exposure should be considered to improve vaccination strategies.

## Data and materials sharing

All data are available from the corresponding authors upon request and will be deposited at the Universitat de Barcelona open repository. The materials generated in this study are available from the corresponding contact with a completed Materials Transfer Agreement.

## Data Availability

All data are available from the corresponding authors upon request and will be deposited at the Universitat de Barcelona open repository.

## Acknowledgments

We thank the volunteers for their participation in the study and the clinicians from IDIAP Jordi Gol for recruitment, sample and data collection. We are very grateful to Ibrahim Abubakar and Adelaida Sarukhan for their intellectual input, and to Jana Kovar for assistance in coordination. Special thanks to Dídac Macià for statistical analysis advice, Alfons Jiménez and Marta Vidal for binding antibody assays, Sara Corral for support during T-cell assay, Diana Barrios for sample processing, and Laura Puyol for logistics coordination.

This work was supported by the European Union under grant agreement no. 101046314 (END-VOC) and by the Fundació Privada Daniel Bravo Andreu. RR had the support of the Health Department, Catalan Government (PERIS SLT017/20/000224). GM was supported by RYC 2020-029886-I/AEI/10.13039/501100011033, co-funded by European Social Fund (ESF). LS is funded by a Rosetrees Trust and Pears Foundation Advancement Fellowship. We acknowledge support from the grant CEX2023-0001290-S funded by MCIN/AEI/ 10.13039/501100011033, and support from the Generalitat de Catalunya through the CERCA Program/Generalitat de Catalunya 2017 SGR 252 (IrsiCaixa) and 1553 (ISGlobal).

## Contributors

GM, CD, LS and FB designed the study. CD and GM supervised the immune assays and data analysis. RR, MC, CT performed the T-cell assay and RA coordinated the lab activities. EP, BT, JB and LMMA designed, performed the neutralization antibody assays and normalized data to obtain ID50. RR, AY, MEZ, CT performed the analysis and interpretation of results. CMP performed database management and analyses. ARM, JVA, and ARC recruited and followed up participants, sample and data collection. RR and MEZ wrote the initial draft and CD, GM, LS, FB reviewed the manuscript. All authors contributed, read and approved the final manuscript.

