## Supplementary data for "Initial antigen encounter determines robust T-cell immunity against SARS-CoV-2 BA.2.86 variant three years later"

Tables S1 and S2

Figures S1 – S10

**Table S1. Immunodominant CD8⁺ T-cell epitopes in SARS-CoV-2 spike protein**

| Epitope number | Epitope number in Grifoni et al. | Epitope Sequence | Domain | BA.2.86 | JN.1 | Other | Length | Antigen | Start | End |
|---|---|---|---|---|---|---|---|---|---|---|
| 1 | 138 | YTNSFTRGVY | NTD | | | | 10 | S | 28 | 37 |
| 2 | 146 | STQDLFLPFF | NTD | **L**TQDLFLPFF | | | 10 | S | 50 | 59 |
| 3 | 154 | RFDNPVLPF | NTD | | | | 9 | S | 78 | 86 |
| 4 | 159 | GVYFASTEK | NTD | | | | 9 | S | 89 | 97 |
| 5 | 160 | TEKSNIIRGW | NTD | | | | 10 | S | 95 | 104 |
| 6 | 162 | TLDSKTQSL | NTD | | | | 9 | S | 109 | 117 |
| 7 | 172 | SSANNCTFEY | NTD | | | | 10 | S | 161 | 170 |
| 8 | 179 | FVFKNIDGY | NTD | | | | 9 | S | 192 | 200 |
| 9 | 182 | IYSKHTPINL | NTD | | | IYSKHTPILV | 10 | S | 203 | 212 |
| 10 | 183 | TPINLVRDL | NTD | TPINLVRD**F** | | TPILVRDLP | 9 | S | 208 | 216 |
| 11 | 185 | LPQGFSAL | NTD | **F**PQGFSAL | | | 8 | S | 216 | 223 |
| 12 | 192 | LLALHRSYL | NTD | LLAL**N**RSYL | | | 9 | S | 241 | 249 |
| 13 | 193 | WTAGAAAYY | NTD | WTAGAA**D**YY | | | 9 | S | 258 | 266 |
| 14 | 200 | YLQPRTFLL | NTD | | | | 9 | S | 269 | 277 |
| 15 | 220 | RISNCVADY | RBD | | | | 9 | S | 357 | 365 |
| 16 | 221 | CVADYSVLY | RBD | | | | 9 | S | 361 | 369 |
| 17 | 225 | KCYGVSPTK | RBD | | | | 9 | S | 378 | 386 |
| 18 | 230 | KIADYNYKL | RBD | | | | 9 | S | 417 | 425 |
| 19 | 231 | KLPDDFTGCV | RBD | | | | 10 | S | 424 | 433 |
| 20 | 232 | NLDSKVGGNY | RBM | NLDSK**HS**GNY | | | 10 | S | 440 | 449 |
| 21 | 237 | NYNYLYRLF | RBM | NY**D**Y**W**YRLF | NYNYLYR**S**F | | 9 | S | 448 | 456 |
| 22 | 247 | YFPLQSYGF | RBM | | | | 9 | S | 489 | 497 |
| 23 | 249 | FQPTNGVGY | RBM | | | | 9 | S | 497 | 505 |
| 24 | 253 | QPYRVVVL | RBM | | | | 8 | S | 506 | 513 |
| 25 | 258 | GPKKSTNLV | RBM | | | | 9 | S | 526 | 534 |
| 26 | 264 | EILDITPCSF | S1 | | | | 10 | S | 583 | 592 |
| 27 | 276 | IGAEHVNNSY | S1 | | | | 10 | S | 651 | 660 |
| 28 | 281 | SPRRARSVA | CS | S**R**RRARSVA | | | 9 | S | 680 | 688 |
| 29 | 284 | SVASQSIIAY | CS | | | | 10 | S | 686 | 695 |
| 30 | 285 | VASQSIIAY | CS | | | | 9 | S | 687 | 695 |
| 31 | 286 | SIIAYTMSL | CS | | | | 9 | S | 691 | 699 |
| 32 | 295 | FTISVTTEIL | S2 | | | | 10 | S | 718 | 727 |
| 33 | 296 | TEILPVSMTK | S2 | | | | 10 | S | 724 | 733 |
| 34 | 300 | TECSNLLLQY | S2 | | | | 10 | S | 747 | 756 |
| 35 | 301 | LLQYGSFCT | S2 | | | | 9 | S | 753 | 761 |
| 36 | 313 | LLFNKVTLA | S2 | | | | 9 | S | 821 | 829 |
| 37 | 320 | LTDEMIAQY | S2 | | | | 9 | S | 865 | 873 |
| 38 | 323 | MIAQYTSAL | S2 | | | | 9 | S | 869 | 877 |
| 39 | 324 | GTITSGWTF | S2 | | | | 9 | S | 880 | 888 |
| 40 | 330 | TQNVLYENQK | S2 | | | | 10 | S | 912 | 921 |
| 41 | 332 | NQKLIANQF | S2 | | | | 9 | S | 919 | 927 |
| 42 | 339 | VLNDILSRL | S2 | | | | 9 | S | 976 | 984 |
| 43 | 341 | RLDKVEAEV | S2 | | | | 9 | S | 983 | 991 |
| 44 | 349 | RLQSLQTYV | S2 | | | | 9 | S | 1000 | 1008 |
| 45 | 355 | YHLMSFPQSA | S2 | | | | 10 | S | 1047 | 1056 |
| 46 | 356 | HLMSFPQSA | S2 | | | | 9 | S | 1048 | 1056 |
| 47 | 362 | APHGVVFLHV | S2 | | | | 10 | S | 1056 | 1065 |
| 48 | 365 | VVFLHVTYV | S2 | | | | 9 | S | 1060 | 1068 |
| 49 | 376 | GTHWFVTQR | S2 | | | | 9 | S | 1099 | 1107 |
| 50 | 386 | RLNEVAKNL | S2 | | | | 9 | S | 1185 | 1193 |
| 51 | 387 | NLNESLIDL | S2 | | | | 9 | S | 1192 | 1200 |

| | | | | | | | |
|---|---|---|---|---|---|---|---|
| 52 | 395 | QYIKWPWYI | S2 | 9 | S | 1208 | 1216 |
| 53 | 396 | KWPWYIWLGF | S2 | 10 | S | 1211 | 1220 |
| 54 | 397 | FIAGLIAIV | S2 | 9 | S | 1220 | 1228 |
| 55 | 400 | SEPVLKGVKL | S2 | 10 | S | 1261 | 1270 |

**Table S2. Immunodominant CD4+ T-cell epitopes in SARS-CoV-2 spike protein**

| Epitope number | Epitope number in Grifoni et al. | Epitope Sequence | Domain | BA.2.86 | JN.1 | Other | Length | Antigen | Start | End |
|---|---|---|---|---|---|---|---|---|---|---|
| 1 | 7 | SVLHSTQDLFLPFFS | NTD | SVLH**L**TQDLFLPFFS | | | 15 | S | 46 | 60 |
| 2 | 17 | NNATNVVIKVCEFQF | NTD | NNATNV**F**IKVCEFQF | | | 15 | S | 121 | 135 |
| 3 | 19 | CEFQFCNDPFLGVYY | NTD | | | | 15 | S | 131 | 145 |
| 4 | 22 | SSANNCTFEYVSQPF | NTD | | | | 15 | S | 161 | 175 |
| 5 | 23 | CTFEYVSQPFLMDLE | NTD | | | | 15 | S | 166 | 180 |
| 6 | 25 | LMDLEGKQGNFKNLR | NTD | | | | 15 | S | 176 | 190 |
| 7 | 27 | EFVFKNIDGYFKIYS | NTD | | | | 15 | S | 191 | 205 |
| 8 | 28 | NIDGYFKIYSKHTPI | NTD | | | | 15 | S | 196 | 210 |
| 9 | 31 | KHTPINLVRDLPQGF | NTD | | | KHTPILVRDLPQGF | 15 | S | 206 | 220 |
| 10 | 33 | NLVRDLPQGFSALEP | NTD | | | LVRDLPQGFSALEP | 15 | S | 211 | 225 |
| 11 | 34 | LPQGFSALEPLVDLP | NTD | **F**PQGFSALEPLVDLP | | | 15 | S | 216 | 230 |
| 12 | 36 | IGINITRFQTLLALH | NTD | IGINITRFQTLLAL**N** | | | 15 | S | 231 | 245 |
| 13 | 37 | ITRFQTLLALHRSYL | NTD | | | | 15 | S | 235 | 249 |
| 14 | 38 | TRFQTLLALHRSYLT | NTD | | | | 15 | S | 236 | 250 |
| 15 | 39 | LLALHRSYLTPGDSS | NTD | LLAL**N**RSYLTPGDSS | | | 15 | S | 241 | 255 |
| 16 | 45 | QPTESIVRFPNITNL | RBD | QPTESIVRFPN**V**TNL | | | 15 | S | 321 | 335 |
| 17 | 46 | IVRFPNITNLCPFGE | RBD | IVRFPN**V**TNLCPFGE | | | 15 | S | 326 | 340 |
| 18 | 47 | CPFGEVFNATRFASV | RBD | | | | 15 | S | 336 | 350 |
| 19 | 48 | VFNATRFASVYAWNR | RBD | | | | 15 | S | 341 | 355 |
| 20 | 49 | RFASVYAWNRKRISN | RBD | RFASVYAWNR**T**RISN | | | 15 | S | 346 | 360 |
| 21 | 61 | GCVIAWNSNNLDSKV | RBD | GCVIAWNSNNLDSK**H** | | | 15 | S | 431 | 445 |
| 22 | 63 | GGNYNYLYRLFRKSN | RBD | GGNY**D**YLYRLFRKSN | GGNYNYLYR**S**FRKSN | | 15 | S | 446 | 460 |
| 23 | 70 | QPYRVVVLSFELLHA | RBD | | | | 15 | S | 506 | 520 |
| 24 | 77 | SIIAYTMSLGAENSV | CS | | | | 15 | S | 691 | 705 |
| 25 | 78 | AENSVAYSNNSIAIP | S2 | | | | 15 | S | 701 | 715 |
| 26 | 80 | SIAIPTNFTISVTTE | S2 | | | | 15 | S | 711 | 725 |
| 27 | 81 | TNFTISVTTEILPVS | S2 | | | | 15 | S | 716 | 730 |
| 28 | 83 | STECSNLLLQYGSFC | S2 | | | | 15 | S | 746 | 760 |
| 29 | 84 | NLLLQYGSFCTQLNR | S2 | | | | 15 | S | 751 | 765 |

| | | | | | | | |
|---|---|---|---|---|---|---|---|
| 30 | 86 | TQLNRALTGIAVEQD | S2 | 15 | S | 761 | 775 |
| 31 | 90 | NFSQILPDPSKPSKR | S2 | 15 | S | 801 | 815 |
| 32 | 92 | KPSKRSFIEDLLFNK | S2 | 15 | S | 811 | 825 |
| 33 | 93 | SFIEDLLFNKVTLAD | S2 | 15 | S | 816 | 830 |
| 34 | 96 | AGFIKQYGDCLGDIA | S2 | 15 | S | 831 | 845 |
| 35 | 101 | FNGLTVLPPLLTDEM | S2 | 15 | S | 855 | 869 |
| 36 | 103 | TDEMIAQYTSALLAG | S2 | 15 | S | 866 | 880 |
| 37 | 108 | IPFAMQMAYRFNGIG | S2 | 15 | S | 896 | 910 |
| 38 | 109 | QMAYRFNGIGVTQNV | S2 | 15 | S | 901 | 915 |
| 39 | 116 | VQIDRLITGRLQSLQ | S2 | 15 | S | 991 | 1005 |
| 40 | 127 | ELDKYFKNHTSPDVD | S2 | 15 | S | 1151 | 1165 |
| 41 | 129 | GINASVVNIQKEIDR | S2 | 15 | S | 1171 | 1185 |

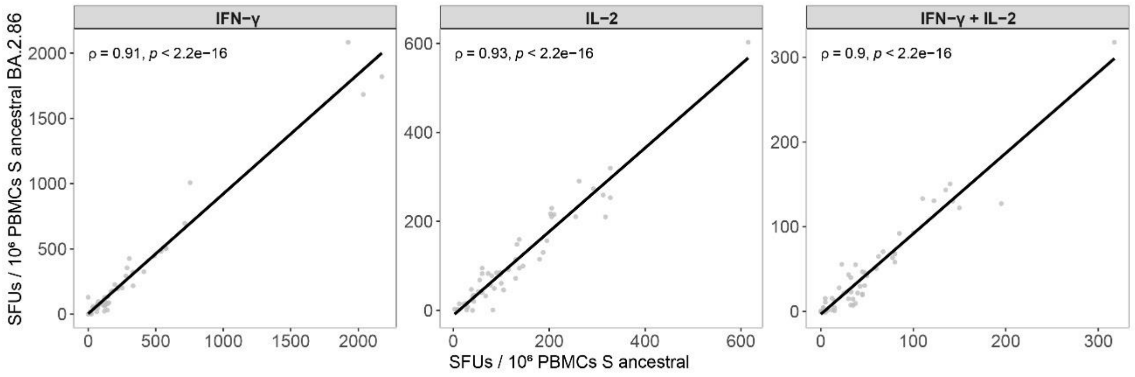


**Figure S1. Correlations of T-cell responses between BA.2.86 and Wuhan.** Spearman's correlation coefficient ρ (Rho) between the magnitude of T-cell responses as SFU / $10^6$ PBMCs of T-cells secreting IFN-γ, IL-2 or IFN-γ + IL-2 (polyfunctional) to ancestral and B.2.86 variant. Interferon-gamma (IFN-γ), interleukin-2 (IL-2), peripheral mononuclear cells (PBMCs), spike (S), spot-forming units (SFU).

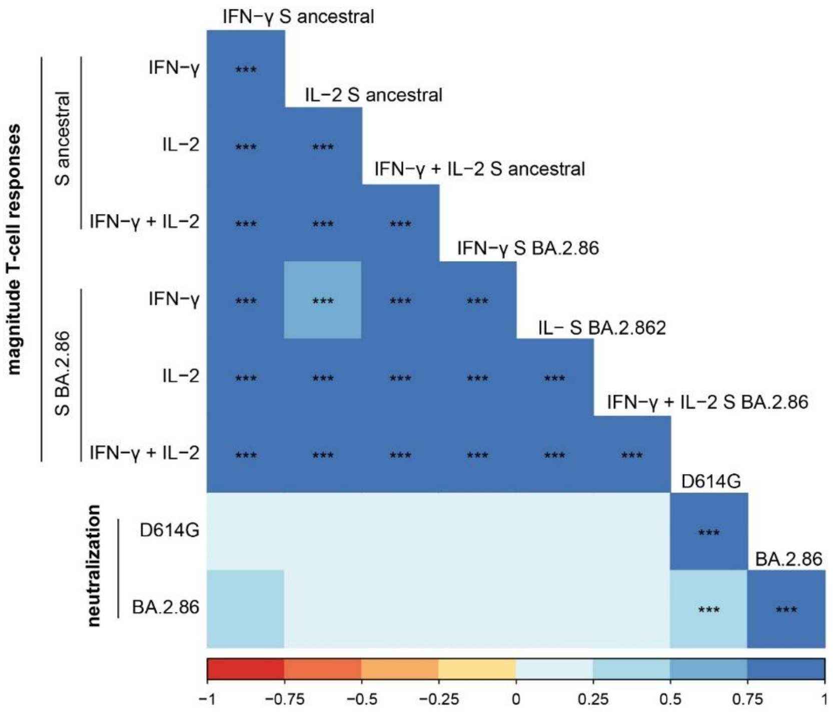


**Figure S2. Correlations of T-cell responses and plasma neutralizing activity.** Heatmap illustrating the Spearman's correlation coefficient ρ (Rho) between the magnitude of T-cell responses as SFU / $10^6$ PBMCs of T-cells secreting IFN-γ, IL-2 or IFN-γ + IL-2 (polyfunctional) and plasma neutralizing activity as ID50. p-values: * ≤ 0.05, ** ≤ 0.01 and *** ≤ 0.001. Interferon-gamma (IFN-γ), interleukin-2 (IL-2), peripheral mononuclear cells (PBMCs), spike (S), receptor binding domain (RBD), spot-forming units (SFU).

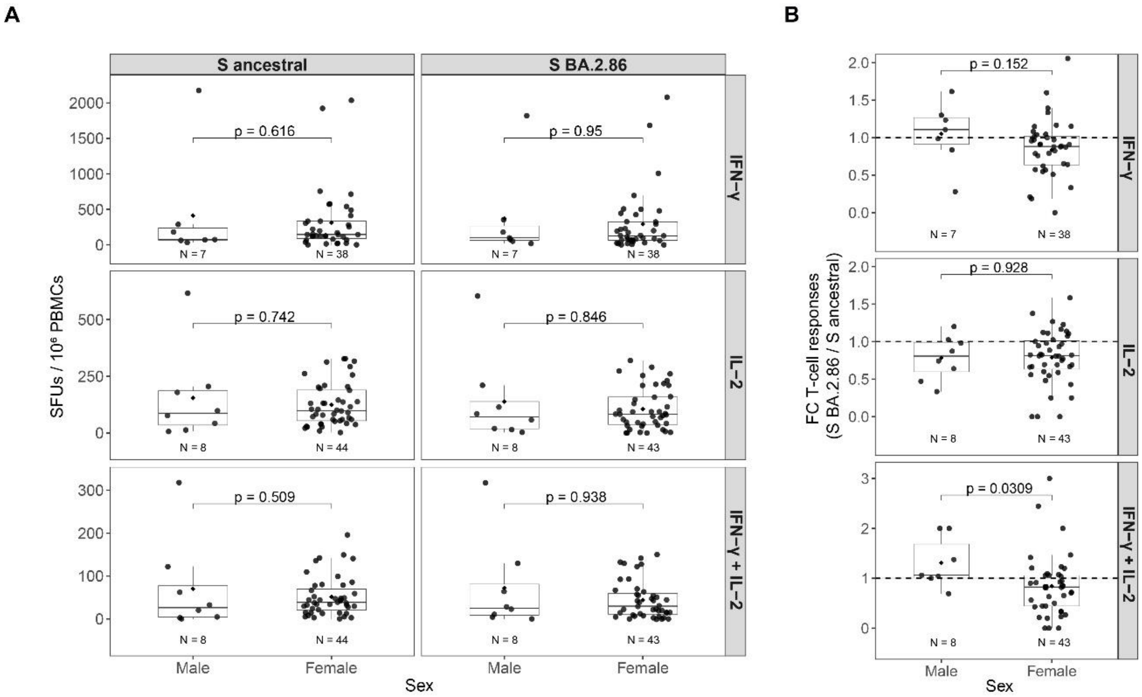


**Figure S3. T-cell responses by sex.** (**A**) Magnitude of T-cell responses as SFU / $10^6$ PBMCs of T-cells secreting IFN-γ, IL-2 or IFN-γ + IL-2 (polyfunctional) and (**B**) BA.2.86 cross-recognition as FC in T-cell responses to BA.2.86 with respect to ancestral strain (BA.2.86 / ancestral) by sex. T-cell responses were compared by Wilcoxon rank-sum test. Boxplots represent median (bold line), the mean (black diamond), $1^{st}$ and $3^{rd}$ quartiles (box), and largest and smallest values within 1.5 times the interquartile range (whiskers). Interferon-gamma (IFN-γ), interleukin-2 (IL-2), peripheral mononuclear cells (PBMCs), spike (S, spot-forming units (SFU).

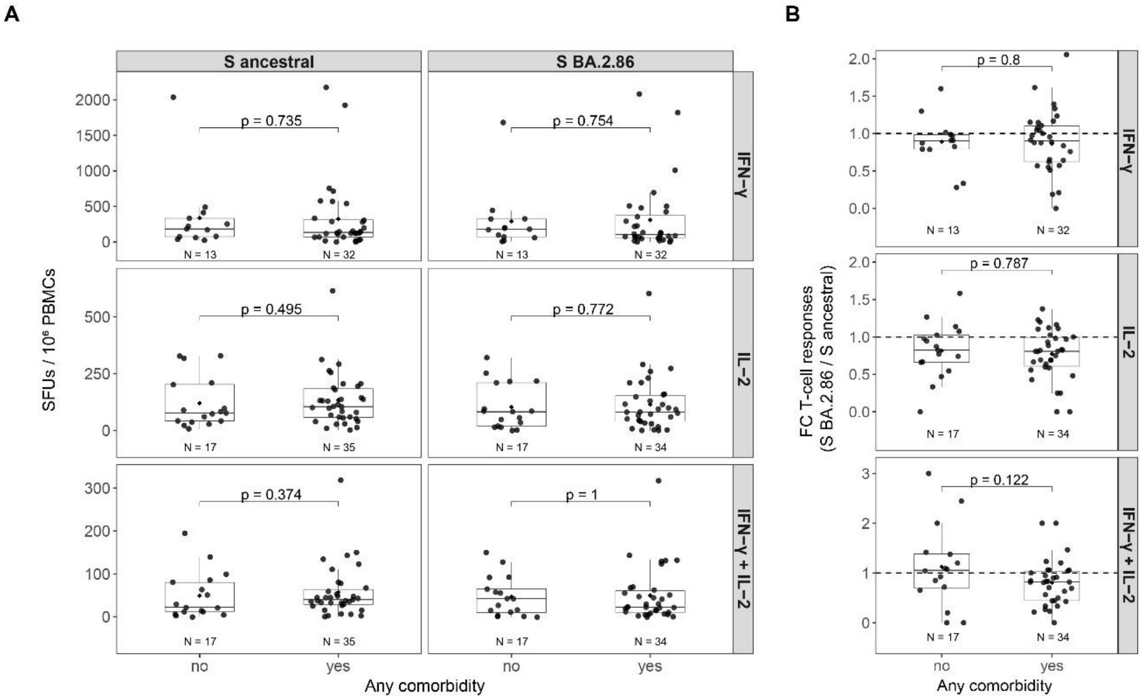


**Figure S4. T-cell responses by any comorbidity.** (**A**) Magnitude of T-cell responses as SFU / $10^6$ PBMCs of T-cells secreting IFN-γ, IL-2 or IFN-γ + IL-2 (polyfunctional) and (**B**) BA.2.86 cross-recognition as FC in T-cell responses to BA.2.86 with respect to ancestral strain (BA.2.86 / ancestral) by any comorbidity. T-cell responses were compared by Wilcoxon rank-sum test. Boxplots represent median (bold line), the mean (black diamond), 1st and 3rd quartiles (box), and largest and smallest values within 1.5 times the interquartile range (whiskers). Any comorbidity includes: Chronic obstructive pulmonary disease, Asthma, Cardiac, Neurologic, Digestive, Chronic kidney disease, Autoimmune, Cancer, Immunosuppression, Mellitus diabetes, Dyslipidaemia, Arterial hypertension, Hypothyroidism, Depression, Obesity and Allergies. Interferon-gamma (IFN-γ), interleukin-2 (IL-2), peripheral mononuclear cells (PBMCs), spike (S, spot-forming units (SFU).

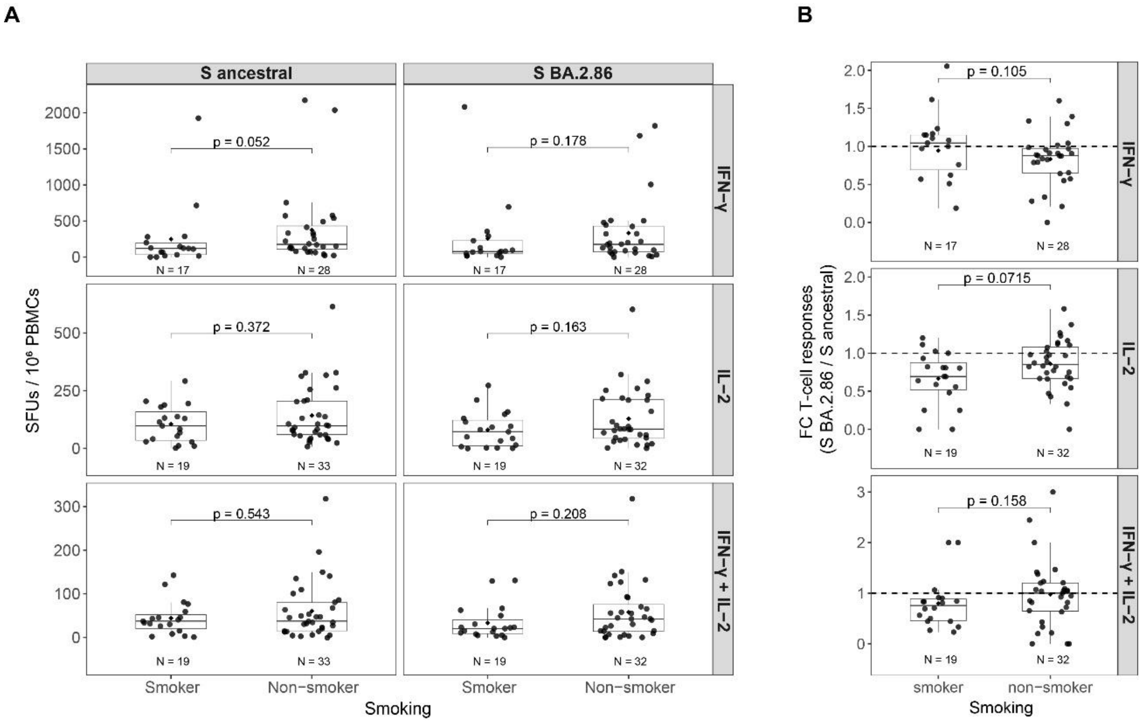


**Figure S5. T-cell responses by smoking.** (**A**) Magnitude of T-cell responses as SFU / $10^6$ PBMCs of T-cells secreting IFN-γ, IL-2 or IFN-γ + IL-2 (polyfunctional) and (**B**) BA.2.86 cross-recognition as FC in T-cell responses to BA.2.86 with respect to ancestral strain (BA.2.86 / ancestral) by smoking. T-cell responses were compared by Wilcoxon rank-sum test. Boxplots represent median (bold line), the mean (black diamond), 1st and 3rd quartiles (box), and largest and smallest values within 1.5 times the interquartile range (whiskers). Interferon-gamma (IFN-γ), interleukin-2 (IL-2), peripheral mononuclear cells (PBMCs), spike (S, spot-forming units (SFU).

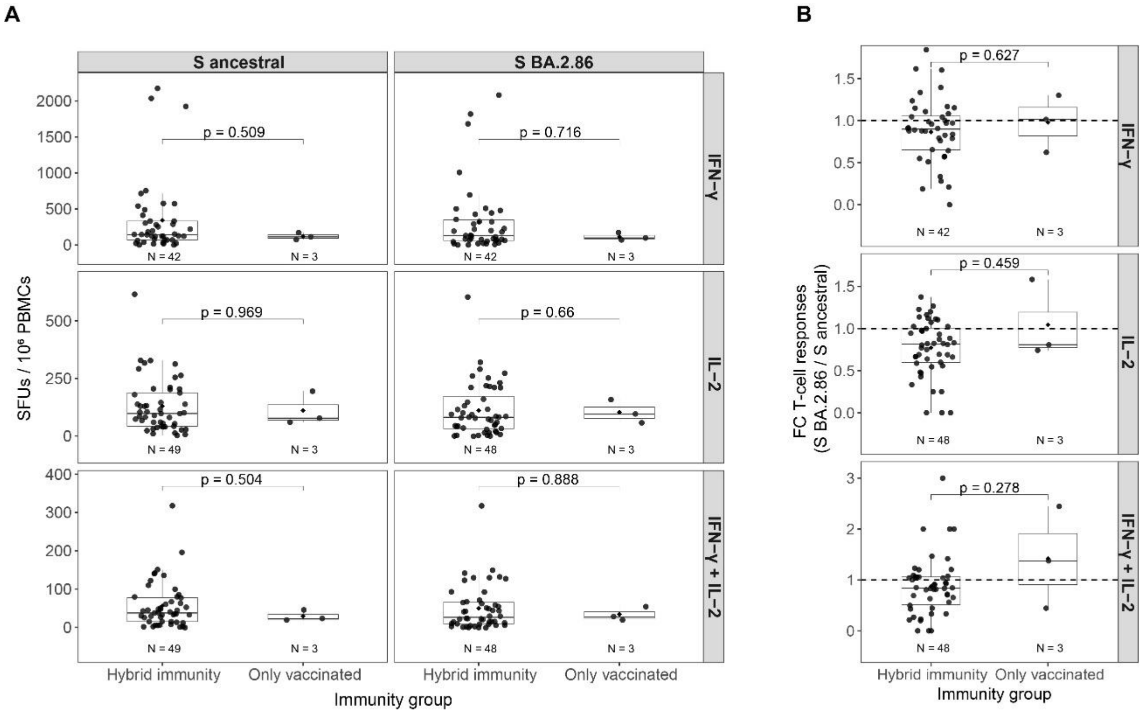


**Figure S6. T-cell responses by immunity groups.** (**A**) Magnitude of T-cell responses as SFU / $10^6$ PBMCs of T-cells secreting IFN-γ, IL-2 or IFN-γ + IL-2 (polyfunctional) and (**B**) BA.2.86 cross-recognition as FC in T-cell responses to BA.2.86 with respect to ancestral strain (BA.2.86 / ancestral) by immunity groups. T-cell responses were compared by Wilcoxon rank-sum test. Boxplots represent median (bold line), the mean (black diamond), $1^{st}$ and $3^{rd}$ quartiles (box), and largest and smallest values within 1.5 times the interquartile range (whiskers). Interferon-gamma (IFN-γ), interleukin-2 (IL-2), peripheral mononuclear cells (PBMCs), spike (S, spot-forming units (SFU).

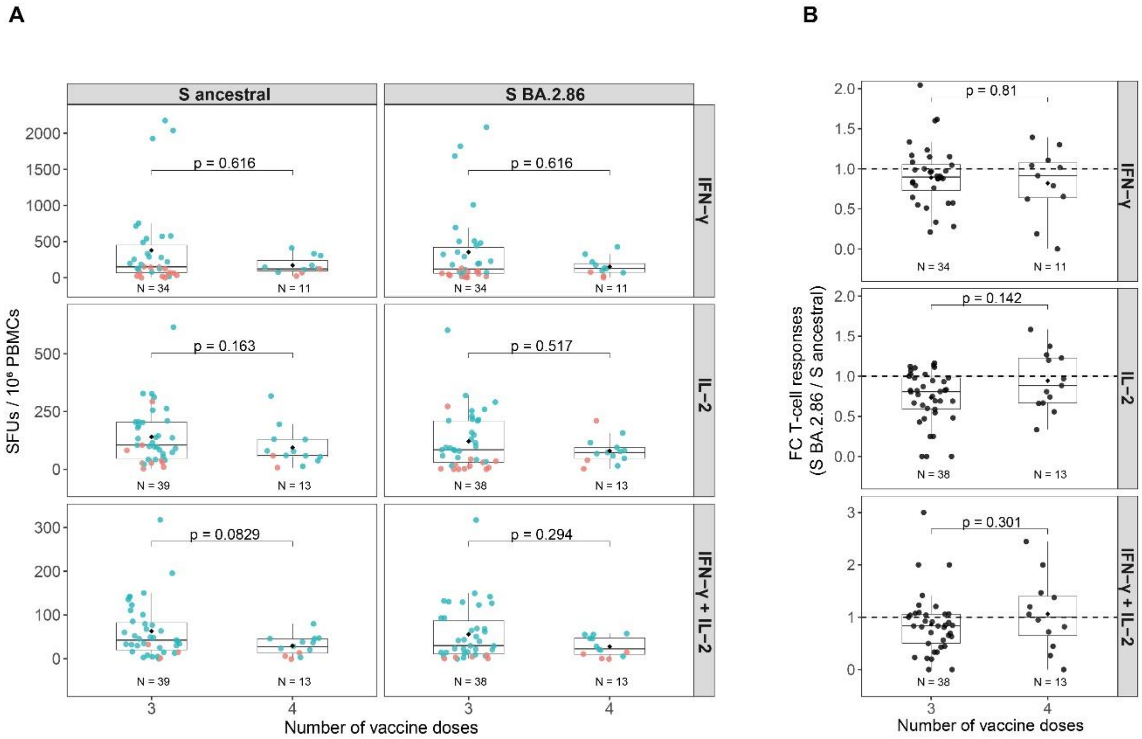


**Figure S7. T-cell responses by number of vaccine doses. A**) Magnitude of T-cell responses as SFU / $10^6$ PBMCs of T-cells secreting IFN-γ, IL-2 or IFN-γ + IL-2 (polyfunctional) and (**B**) BA.2.86 cross-recognition as FC in T-cell responses to BA.2.86 with respect to ancestral strain (BA.2.86 / ancestral) by number of vaccine doses. T-cell responses were compared by Wilcoxon rank-sum test. Boxplots represent median (bold line), the mean (black diamond), 1st and 3rd quartiles (box), and largest and smallest values within 1.5 times the interquartile range (whiskers). Interferon-gamma (IFN-γ), interleukin-2 (IL-2), peripheral mononuclear cells (PBMCs), spike (S, spot-forming units (SFU).

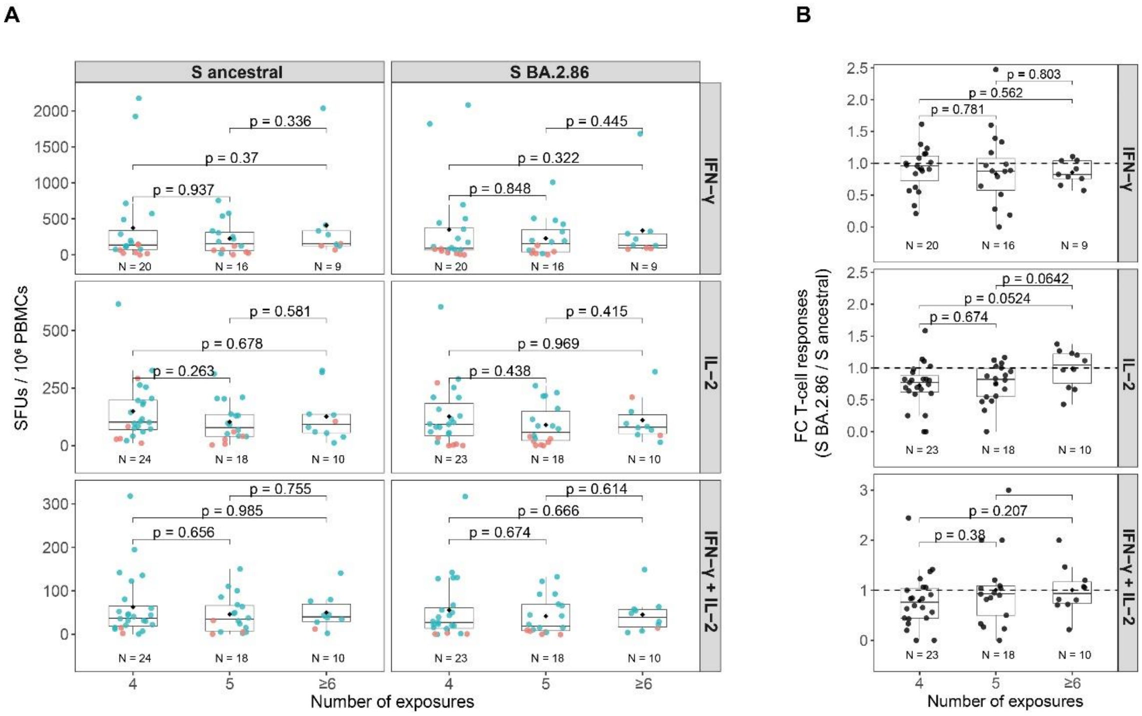


**Figure S8. T-cell responses by number of total exposures. A**) Magnitude of T-cell responses as SFU / $10^6$ PBMCs of T-cells secreting IFN-γ, IL-2 or IFN-γ + IL-2 (polyfunctional) and (**B**) BA.2.86 cross-recognition as FC in T-cell responses to BA.2.86 with respect to ancestral strain (BA.2.86 / ancestral) by number of total exposures (vaccine doses and infections). T-cell responses were compared by Wilcoxon rank-sum test. Boxplots represent median (bold line), the mean (black diamond), 1st and 3rd quartiles (box), and largest and smallest values within 1.5 times the interquartile range (whiskers). Interferon-gamma (IFN-γ), interleukin-2 (IL-2), peripheral mononuclear cells (PBMCs), spike (S, spot-forming units (SFU).

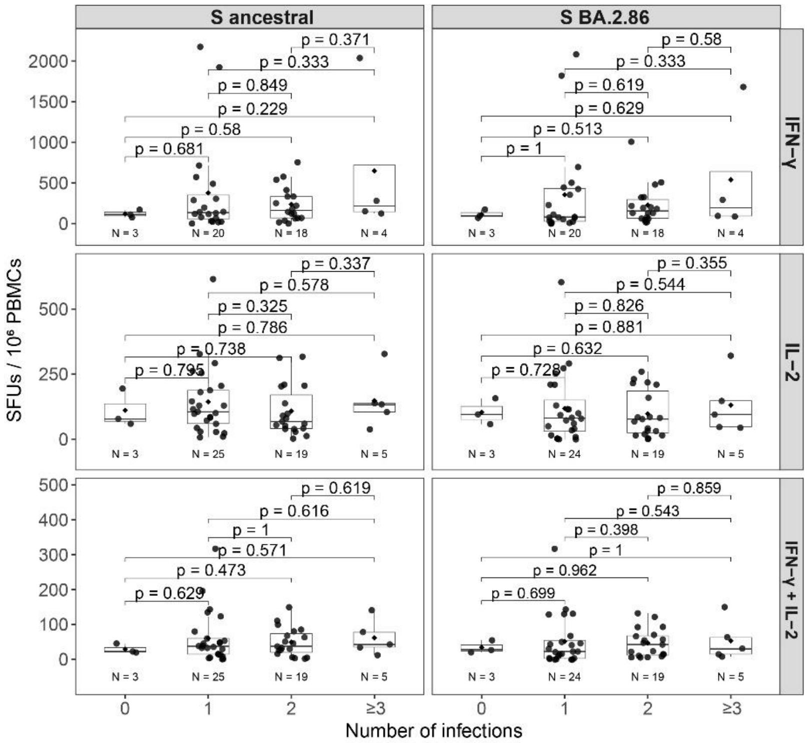


**Figure S9. T-cell responses by number of infections. A**) Magnitude of T-cell responses as SFU / $10^6$ PBMCs of T-cells secreting IFN-γ, IL-2 or IFN-γ + IL-2 (polyfunctional) and (**B**) BA.2.86 cross-recognition as FC in T-cell responses to BA.2.86 with respect to ancestral strain (BA.2.86 / ancestral) by number of infections. T-cell responses were compared by Wilcoxon rank-sum test. Boxplots represent median (bold line), the mean (black diamond), 1st and 3rd quartiles (box), and largest and smallest values within 1.5 times the interquartile range (whiskers). Interferon-gamma (IFN-γ), interleukin-2 (IL-2), peripheral mononuclear cells (PBMCs), spike (S), spot-forming units (SFU).

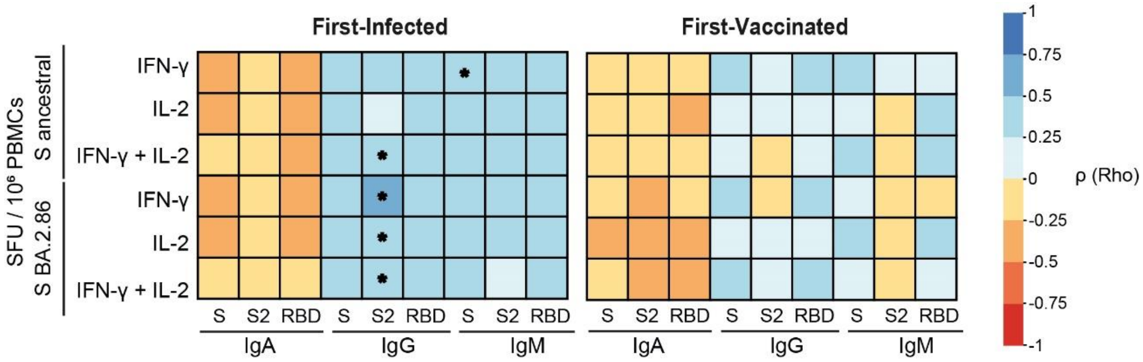


**Figure S10. Correlations of T-cell responses with plasma antibody levels by first antigen encounter groups.** Heatmaps illustrating the Spearman's correlation coefficient ρ (Rho) between the as SFU / $10^6$ PBMCs of T-cells secreting IFN-γ, IL-2 or IFN-γ + IL-2 (polyfunctional) with the antibody responses (median fluorescence intensity (MFI) of IgA, IgG, and IgM) to S, S2 and RBD from ancestral strain. p-values: * ≤ 0.05, ** ≤ 0.01 and *** ≤ 0.001. Interferon-gamma (IFN-γ), interleukin-2 (IL-2), peripheral mononuclear cells (PBMCs), spike (S), receptor binding domain (RBD), spot-forming units (SFU).
